# Utilizing ChatGPT to select literature for meta-analysis shows workload reduction while maintaining a similar recall level as manual curation

**DOI:** 10.1101/2023.09.06.23295072

**Authors:** Xiangming Cai, Yuanming Geng, Yiming Du, Bart Westerman, Duolao Wang, Chiyuan Ma, Juan J. Garcia Vallejo

**Affiliations:** Department of Molecular Cell Biology & Immunology, Amsterdam Infection & Immunity Institute and Cancer Center Amsterdam, Amsterdam UMC, Vrije Universiteit Amsterdam, Amsterdam, The Netherlands; Department of Neurosurgery, Jinling Hospital, Nanjing, China; Department of Neurosurgery, Affiliated Jingling Hospital, Nanjing Medical University, Nanjing, China; Department of System Engineering and Engineering Management, The Chinese University of Hong Kong, Hong Kong, China; Department of Neurosurgery, Cancer Center Amsterdam, Brain tumor center Amsterdam, Amsterdam UMC location Vrije Universiteit Amsterdam, Amsterdam, The Netherlands; Department of Clinical Sciences, Liverpool School of Tropical Medicine, Liverpool, United Kingdom; School of Medicine, Southeast University, Nanjing, China; Department of Neurosurgery, Affiliated Jinling Hospital, Medical School of Nanjing University, Nanjing, China; Department of Neurosurgery, Jinling Hospital, the First School of Clinical Medicine, Southern Medical University, Nanjing, China

**Keywords:** Large language model, ChatGPT, Meta-analysis, Glioma, Immunotherapy

## Abstract

**Background:** Large language models (LLMs) like ChatGPT showed great potential in aiding medical research. A heavy workload in filtering records is needed during the research process of evidence-based medicine, especially meta-analysis. However, no study tried to use LLMs to help screen records in meta-analysis.

**Objective:** In this research, we aimed to explore the possibility of incorporating ChatGPT to facilitate the screening step based on the title and abstract of records during meta-analysis.

**Methods:** To assess our strategy, we selected three meta-analyses from the literature, together with a glioma meta-analysis embedded in the study, as additional validation. For the automatic selection of records from curated meta-analyses, a four-step strategy called LARS-GPT was developed, consisting of (1) criteria selection and single-prompt (prompt with one criterion) creation, (2) best combination identification, (3) combined-prompt (prompt with one or more criteria) creation, and (4) request sending and answer summary. Recall, workload reduction, precision, and F1 score were calculated to assess the performance of LARS-GPT.

**Results:** A variable performance was found between different single-prompts with a mean recall of 0.841. Based on these single-prompts, we were able to find combinations with performance better than the pre-set threshold. Finally, with a best combination of criteria identified, LARS-GPT showed a 39.5% workload reduction on average with a recall greater than 0.9.

**Conclusions:** We show here the groundbreaking finding that automatic selection of literature for meta-analysis is possible with ChatGPT. We provide it here as a pipeline, LARS-GPT, which showed a great workload reduction while maintaining a pre-set recall.

## Introduction

The medical understanding of diseases has advanced rapidly during the last decades, but the translation from bench to bedside is lagging.^1^ Evidence-based medicine (EBM), especially meta-analysis, facilitates the application of novel therapies into clinics; however, the processes of conducting meta-analysis are time-consuming and work intensive.^2^ Artificial intelligence (AI) is becoming ubiquitous in medicine.^1^ And AI-based solutions are developed to reduce human efforts spent on EBM with promising performance.^3^ AI models can provide predicted probability for all records based on “similarity” between them. However, human annotators are needed to train the AI models.^4,5^ What’s more, although it helps accelerating the research process, researchers still need to screen all records.

Recent releases of large language models (LLMs) like ChatGPT have dramatic implications on medical research;^6-8^ however, few studies have evaluated its application in aiding EBM and review writing. Shaib *et al*. utilized ChatGPT (text-davinci-003) to synthesize medical evidence,^9^ and Shuai *et al*. explored its effectiveness in generating Boolean queries for a literature search.^10^ However, no study has investigated its application in compensating or substituting human effort spent on filtering records during meta-analysis, a key issue because of the exponentially increased number of primary literature and systemic reviews required by medical researchers nowadays.^11^

In this study, we aimed to explore the possibility of using ChatGPT to aid the automatic selection of literature records (based on their title and abstract) for meta-analysis by developing a pipeline named LARS-GPT (Literature Records Screener based on ChatGPT). With this study, we show a way to integrate LLMs into the field of EBM, which may impact the research pattern of meta-analysis.

## Methods

### Screen pipeline incorporating ChatGPT: LARS-GPT

In general, the workflow of meta-analysis has the following steps: (1) define research question; (2) select literature databases and design search strategy; (3) screen records based on their titles and abstracts; (4) screen records based on full text of records; (5) extract and synthesis data. In the present study, we focused on incorporating ChatGPT into the third step of this workflow.

To do that, we designed the four-step pipeline, LARS-GPT (**Fig 1**). First, users need to select criteria (some suitable criteria from filtering criteria of meta-analysis) and create a prompt for each criterion (single-prompt; **Table 1**). Second, users need to evaluate these single-prompts using a few records and then select the best combination of single-prompts. Third, users need to choose a prompt strategy and merge single-prompts in the best combination to make a combined prompt (combined-prompt; **Supplementary File 1**) in accordance with the selected prompt strategy. Finally, the combined-prompt, together with the title and abstract of each record, will be submitted to ChatGPT as chat completion. The decisions about whether a record meets the user’s criteria will then be extracted from returned answers. In this study, we evaluated both GPT-3.5 (gpt-3.5-turbo-0301) and GPT-4 (gpt-4-0314) using the API (Application Programming Interface) provided by OpenAI. In practice, LARS-GPT could be performed in batches using Python.

**Table 1.**
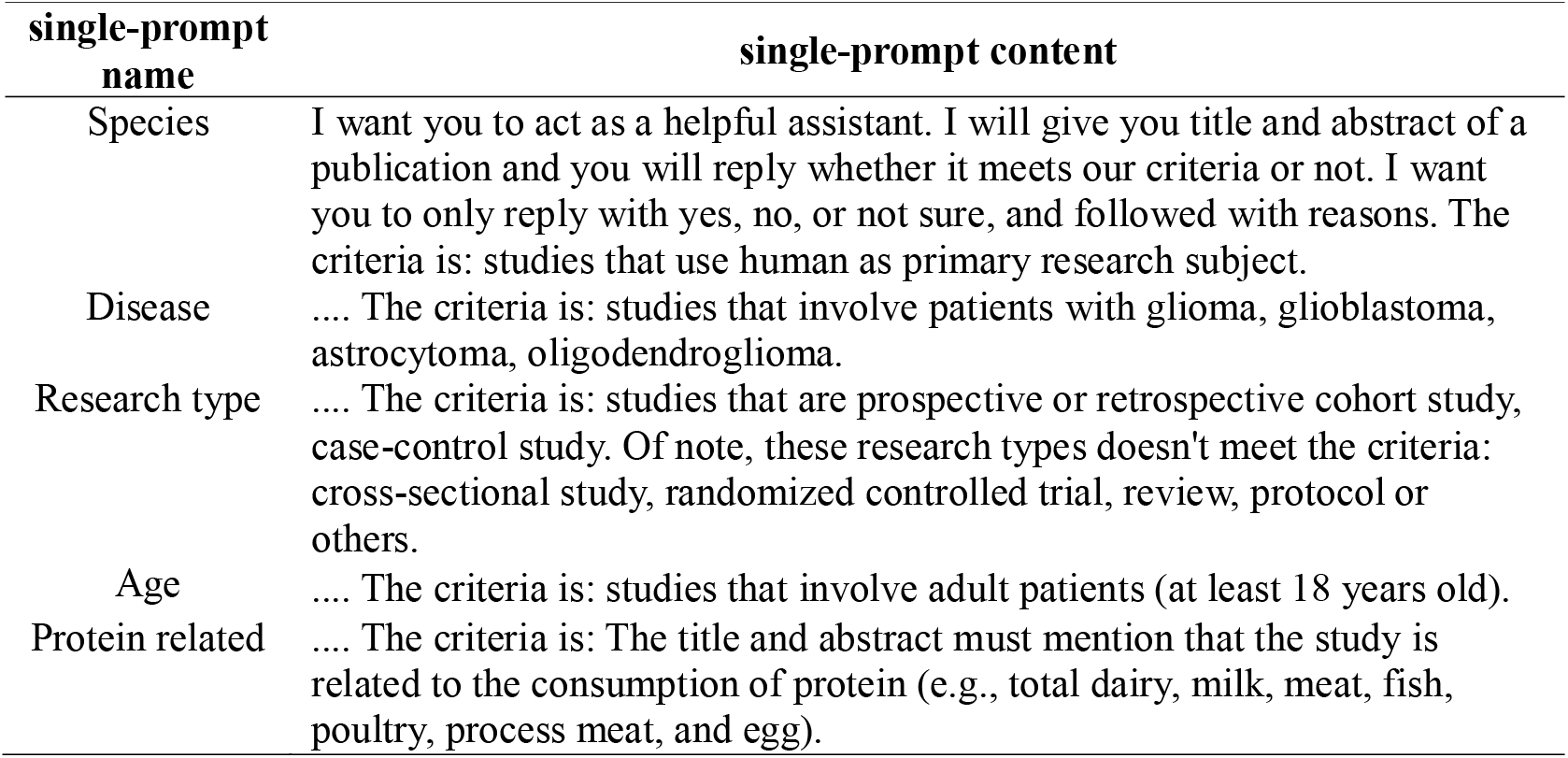
Representative prompt with single criterion (single-prompt)

| single-prompt name | single-prompt content |
| --- | --- |
| Species | I want you to act as a helpful assistant. I will give you title and abstract of a publication and you will reply whether it meets our criteria or not. I want you to only reply with yes, no, or not sure, and followed with reasons. The criteria is: studies that use human as primary research subject. |
| Disease | .... The criteria is: studies that involve patients with glioma, glioblastoma, astrocytoma, oligodendroglioma. |
| Research type | .... The criteria is: studies that are prospective or retrospective cohort study, case-control study. Of note, these research types doesn't meet the criteria: cross-sectional study, randomized controlled trial, review, protocol or others. |
| Age | .... The criteria is: studies that involve adult patients (at least 18 years old). |
| Protein related | .... The criteria is: The title and abstract must mention that the study is related to the consumption of protein (e.g., total dairy, milk, meat, fish, poultry, process meat, and egg). |

**Figure 1.**
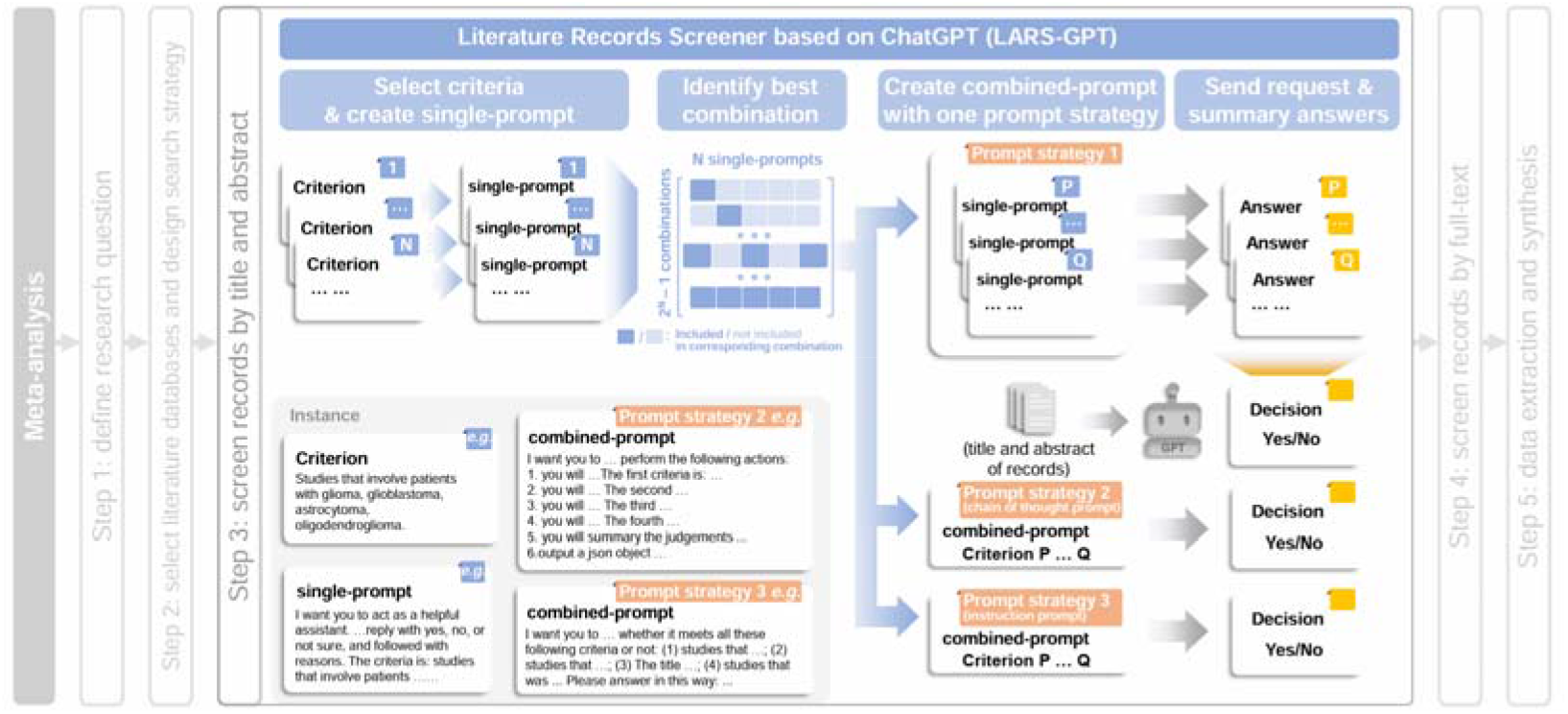
Schematic illustration of the LARS-GPT pipeline Single-prompt represents a prompt with only one criterion. Combined-prompt stands for the prompt with more than one criterion. Color of labels: single-prompt (blue), combined-prompt and prompt strategy (orange), answer and decision (yellow), and true outcome of validation datasets (green).

### Selection of validation meta-analyses

To cover broad medical fields, we selected three high-quality published meta-analyses as validation datasets, which focused on inflammatory bowel diseases (IBD),^12^ diabetes mellitus (DM),^13^ and sarcopenia,^14^ respectively (**Table 2**). These published meta-analyses provided clear search strategies for Medline/PubMed database and complete list of records that remained after screening based on their titles and abstracts. Thanks to this, we were able to repeat their literature search in Medline/PubMed and match record list to obtain the correct answer that whether these identified records could pass the screening step in a real-world practice (**Table 2; Supplementary File 2**). On top of these published meta-analyses, we conducted a new meta-analysis about glioma. The protocol of the glioma meta-analysis was registered on PROSPERO (CRD42023425790). In doing so, we can evaluate the performance of ChatGPT in a first-hand practice.

**Table 2.**
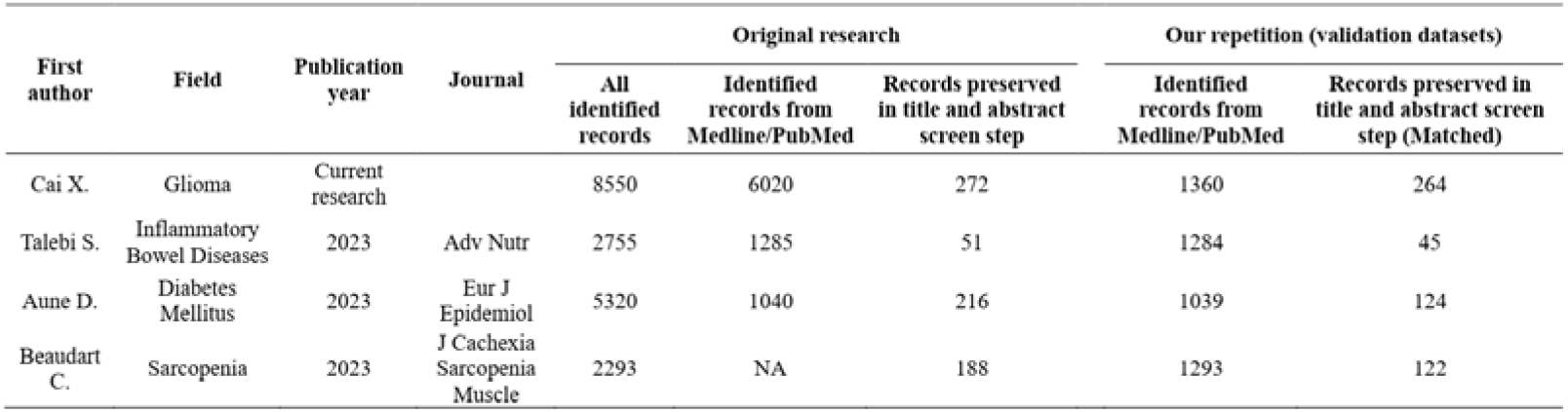
Summary of meta-analyses included as validation datasets for LARS-GPT.

The number of records used for each step evaluation is different, due to the requirements of each step, the workload, and the cost of money. In the final step evaluation with the combined-prompt, almost all records were used for the GPT-3.5 evaluation. However, only 100 randomly selected records were used for the GPT-4 evaluation, due to the limited funding. The detailed randomization method used here can be found in **Supplementary File 3**.

#### Step1: Prompt strategy design

We designed prompts (**Table 1**; **Supplementary File 1**) with the guidance from OpenAI (https://platform.openai.com/docs/guides/gpt-best-practices). However, the high flexibility of prompt and the “black box” nature of ChatGPT made it impossible to design a “best” prompt. In this study, we designed three distinct types of prompt strategies to help create better combined-prompt (**Fig 1 and 2; Supplementary File 1**). For the “single criterion” prompt (prompt strategy 1), we simply maintain these single-prompts in the best combination. ChatGPT will respond to each single-prompt and determine whether a record meets each criterion or not. After receiving answers from ChatGPT, users need to summarize answers for each single-prompt and make a final decision for each record. In this study, as long as there is one answer that is “No”, the final decision for a record is “No”. Otherwise, the final decision will be “Yes”. For the “instruction prompt” (prompt strategy 3) and “chain of thought prompt” (prompt strategy 2), the best combination of single-prompts was merged into one combined-prompt (**Fig 1 and 2; Supplementary File 1**). Users expect a final judgement from ChatGPT directly. In this research, we selected 4-5 criteria from each meta-analysis (**Table 1**; **Supplementary File 1**).

**Figure 2.**
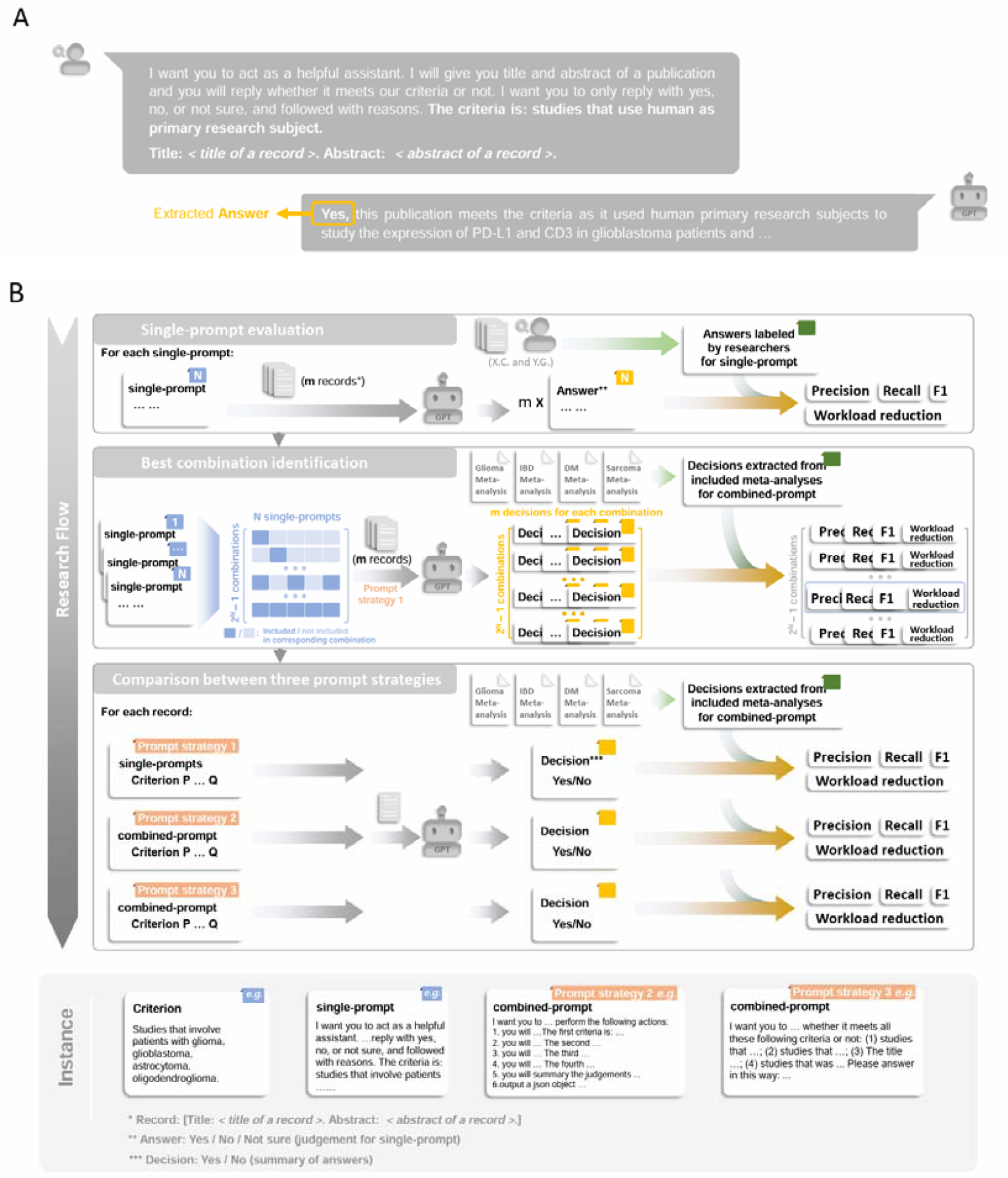
The research flow of this study A representative case showing a request containing a single-prompt and the response from ChatGPT (**A**). The schematic illustrations of the research flow (**B**). Here also shows the detailed input (made by human researchers) for ChatGPT performance metrics calculation. Single-prompt represents a prompt with only one criterion. Combined-prompt stands for the prompt with more than one criterion. Color of labels: single-prompt (blue), combined-prompt and prompt strategy (orange), answer and decision (yellow), and true outcome of validation datasets (green).

#### Step2: Evaluation of the classification performance of single-prompt

We (XC and YG) manually labeled correct answers of each single-prompt within 100 randomly selected records (about 10 positive records and 90 negative records) for each validation meta-analysis. Here, records were called “positive” records if they were remained after the screening step based on their titles and abstracts. Otherwise, they were called “negative” records. To avoid potential bias from the researchers, these records were manually labeled before we tested them on the ChatGPT. With these 100-reords datasets, we evaluated the performance of ChatGPT and a random classifier regarding single-prompts.

#### Step3: Evaluation of single-prompt combination and identification of best combination

Before conducting any evaluation, the “best” combination of single-prompts was unknown. In other words, how many single-prompts and which single-prompts should be selected for combined-prompt creation? To address this question, we evaluated all possible combinations of designed single-prompts. Among these combinations, we selected the best combination, which has a recall ≥ 0.9 and the best workload reduction.

### Statistical analysis

Because of the nature of LLMs, the generated answer from LLMs varies each time, even with exactly identical input. So, we assessed the robustness score of each single-prompt with repeated requests before testing the LARS-GPT pipeline (see **Supplementary File 3**). In general, the returns were stable, with a robustness score ranging from 0.747 to 0.996 (**Supplementary Fig 1 and 2**).

The performance of ChatGPT was assessed with precision, recall, F1 score, and workload reduction metrics. The workload reduction indicator was defined as:

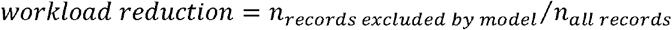

Where n is the number of records. The workload reduction indicator varies between 0 and 1, where 0 indicates none work was reduced and 1 signifies that all work was reduced. For meta-analysis, recall is the most significant indicator, followed with workload reduction, F1, and precision. Throughout the study, we placed greater emphasis on recall and workload reduction as the primary performance metrics.

For other machine learning (ML) models, it’s possible to reach a 100% recall with the compromise of low accuracy. However, due to the distinct mechanisms behind LLMs and other ML models, this is impossible for LLMs-based solutions, at least for our LARS-GPT pipeline. So, in this study, a random classifier was used as a baseline reference (see **Supplementary File 3**). The classifications made by human researchers are used as “true decisions” to calculate the performance metrics of the LARS-GPT pipeline.

## Results

### Single-prompts exhibit distinct performance

The performance of each single-prompt based on GPT-3.5 or GPT-4 was assessed (**Table 3; Supplementary Table 1**). Overall, the majority of prompts had better performance with GPT than a random classifier. The mean recall for GPT was 0.841, with 69.4% single-prompts having a recall higher than 0.8. The GPT-3.5 (mean recall: 0.867) and GPT-4 (mean recall: 0.815) had similar and good recalls. Surprisingly, the recalls could be quite different between these two versions of GPT, even for the same single-prompt, *e*.*g*., the “Control” single-prompt from sarcopenia meta-analysis (GPT-3.5: 0.838; GPT-4: 0.235; **Supplementary Table 1**) and the “Protein related” single-prompt from IBD meta-analysis (GPT-3.5: 0.897; GPT-4: 0.483; **Supplementary Table 1**).

**Table 3.**
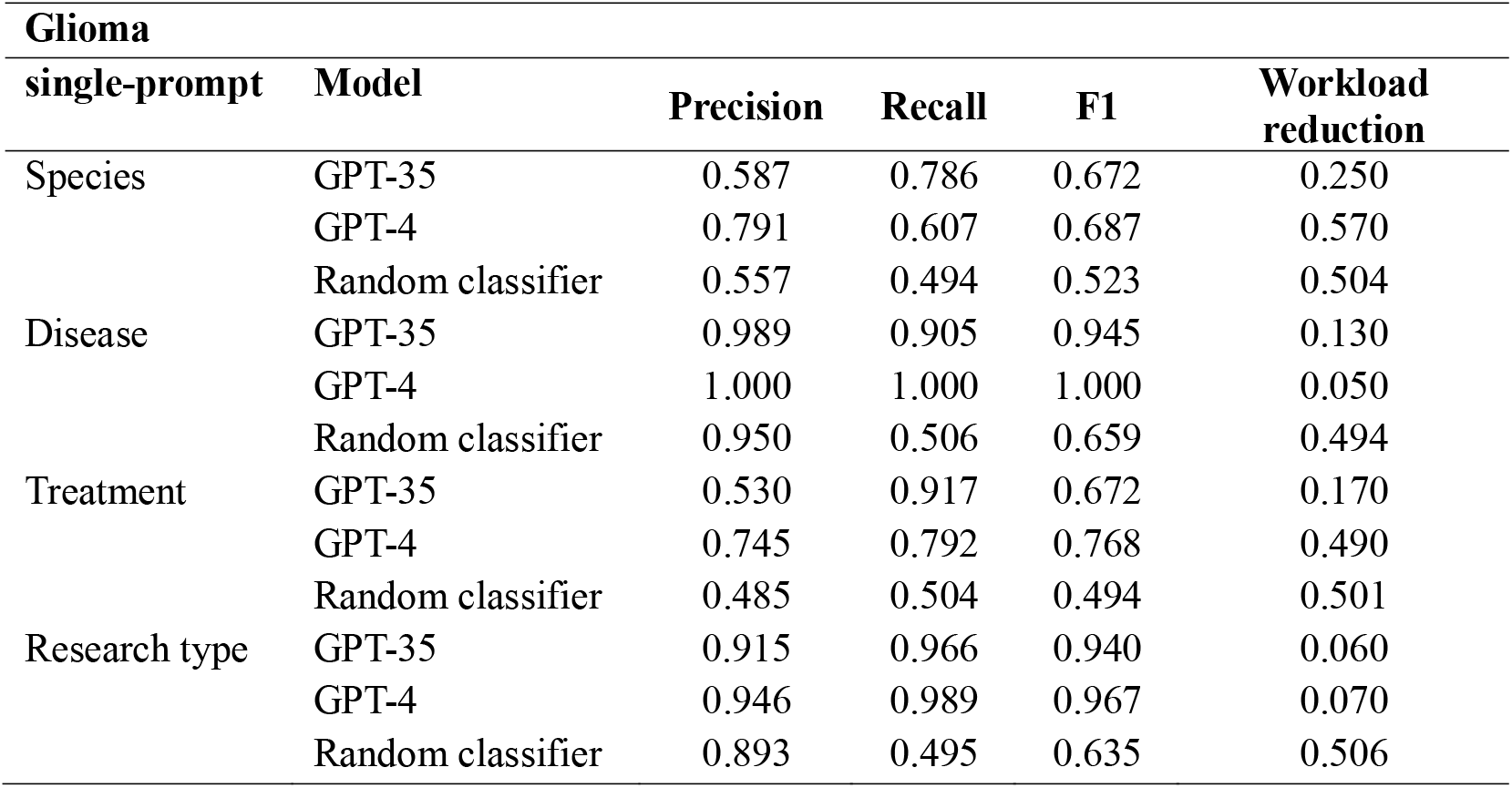
Performance of single-prompts from glioma meta-analysis using GPT-3.5, GPT-4, and random classifier.

Different single-prompts also exhibited distinct recalls. Most single-prompts performed well like the “Research type” prompt from glioma meta-analysis (GPT-3.5: 0.966; GPT-4: 0.989; **Table 3**). However, few single-prompts demonstrated low recalls, *e*.*g*., the “Disease_dm” prompt from IBD meta-analysis (GPT-3.5: 0.554; GPT-4: 0.770; **Supplementary Table 1**).

### The best combination of single-prompts is identified by evaluating the performance of all possible combinations

All combinations of single-prompts were shown in the form of UpSet plots (**Fig 3 and Supplementary Fig 3-5**). As expected, when the number of single-prompts increases, the recall tends to decrease, while workload reduction and precision increase. In general, most combinations presented superior performance compared to a random classifier. To our surprise, it’s not uncommon to find a combination with three single-prompts having a recall of 0.9 or higher, although these cases are all based on GPT-4 (**Fig 3F**; **Supplementary Fig 3F; Supplementary Fig 5F**).

**Figure 3.**
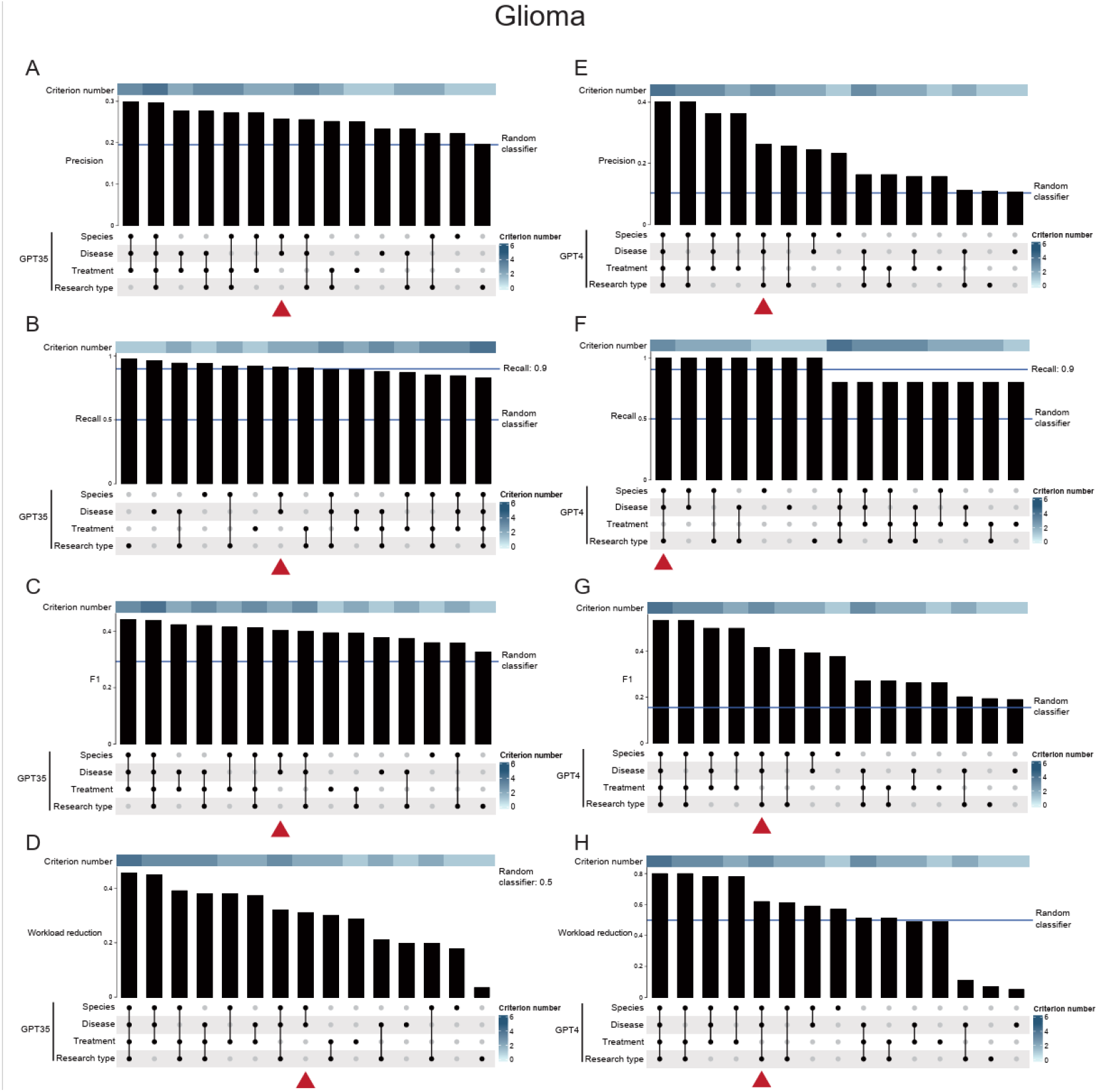
Best combination of single-prompts was identified by evaluating the performance of all combinations of single-prompts from glioma meta-analysis In the Upset plot, the bar chart above represents the evaluation metrics. The dotted line at the bottom presents the single-prompts included in the corresponding combination. Precision (**A, E**), recall (**B, F**), F1 score (**C, G**), and workload reduction (**D, H**) are presented for GPT-3.5 (**A-D**) and GPT-4 (**E-F**), respectively. Best combination is marked with a triangle.

Based on the preset threshold, we identified the best combination with the highest workload reduction from combinations, which have a recall greater than 0.9. However, in the DM meta-analysis using GPT-3.5, there was only one combination with a recall ≥ 0.9, which only included one single-prompt. Because we wanted to evaluate the performance of prompt strategy 2 and 3, which were specifically tailored for combinations involving multiple single-prompts, we selected another combination (“Research type” and “Disease_p”) instead as a sub-best combination for following analyses.

### Three prompt strategies show similar performance

Full combination (including all designed single-prompts) and best combination were both evaluated with three prompt strategies (**Table 4; Supplementary File 1**). Obviously, the best combinations had ideal and much better recalls than full combinations and random classifier. The best combinations demonstrated remarkable recalls ranged from 0.900 to 1.000. The corresponding workload reductions varied from 0.122 to 0.640, with an average of 0.395. The sub-best combination from DM meta-analysis also showed good performance, with recalls ranging from 0.778 to 1.000 and workload reductions varying from 0.280 to 0.460.

**Table 4.**
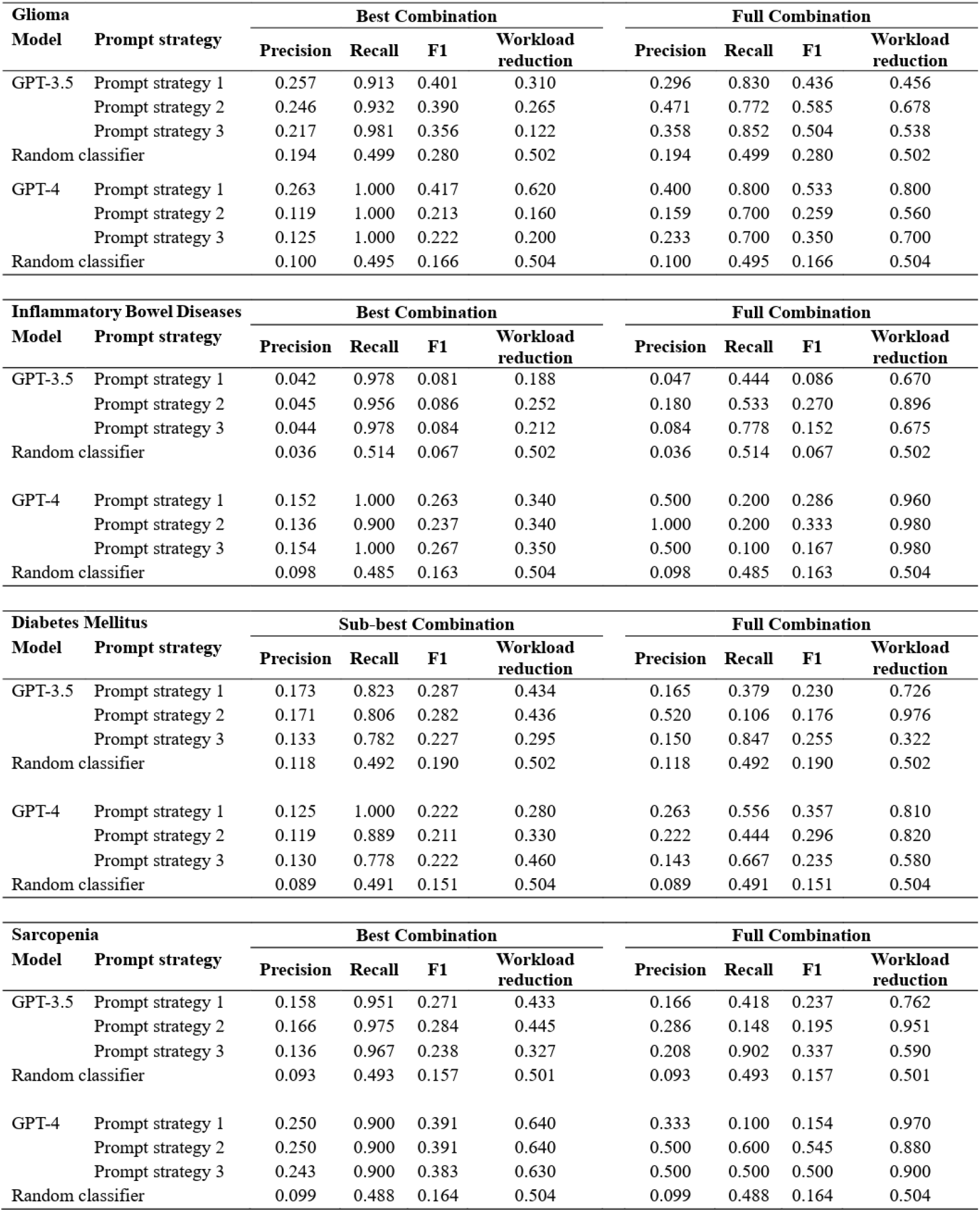
Performance of three prompt strategies with the best combination and the full combination.

These three prompt strategies showed comparable levels of performance (**Supplementary Fig 6A-D**), regarding all four metrics. GPT-3.5 and GPT-4 had similar recalls, precisions, and F1 scores (**Supplementary Fig 6E-G**). However, the workload reduction were slightly higher when using GPT-4 than GPT-3.5 (GPT-3.5 vs. GPT-4, medium: 0.303 vs. 0.345; ANOVA test, P = 0.037; **Supplementary Fig 6H**).

## Discussion

In this research, we developed LARS-GPT and proved that it can greatly reduce the filtering workload while maintaining an ideal recall during the screening step based on the titles and abstracts of records for meta-analysis.

The mechanism employed by LLMs is different from that of previous AI models. Previous AI models applied active learning to select the training dataset and returned all records ordered by a “similarity” index.^5^ However, LLMs have been trained to predict text that follows the input text. By doing so, LLMs can directly answer questions and return whether an input record meet provided criteria or not. Due to the distinct mechanisms applied, previous AI models can reach a perfect recall with the compromise of low accuracy, but not for LLMs-based methods. Thus, we excluded previous AI models as baseline references for performance evaluation in this study.

LLMs have advantages over previous AI models. One advantage is that extra training is unnecessary when applying LLMs to a new meta-analysis (although fine-tuning is possible) because LLMs were pre-trained on large-scale datasets. In comparison, a training dataset is required for every new meta-analysis if choosing previous AI models. Additionally, users do not need to worry about the imbalanced data problem^5^ when using LARS-GPT for the same reason.

An obvious benefit of LARS-GPT is that it could be easily adapted to other LLMs by simply changing the API since most LLMs work similarly. However, the performance of LARS-GPT depends on the performance of the LLM used, which is not guaranteed. We also believe that a well-performed prompt in ChatGPT could be used for other LLMs. However, further research is needed to verify this idea of adapting LARS-GPT to other LLMs.

LLM hallucinations are one issue that has been emphasized in research. These hallucinations occur when a LLM make up fake information and describe it like it is real.^16,17^ LARS-GPT avoids this issue because users need to provide the titles and abstracts of records to ChatGPT, rather than having ChatGPT search for the information. Nonetheless, we did observe instances where ChatGPT made false causal inferences. For example, ChatGPT might give a reason supporting a record meeting one filtering criterion, which is then followed by an opposite judgment. A similar false conclusion may occur when users ask ChatGPT to summarize a final judgment, *e*.*g*. “The publication meets criterion 1, but not criterion 2. So, the publication meets all your criteria”. Despite occasional false judgment, LARS-GPT demonstrated an ideal performance in the current research.

Surprisingly, in this study, GPT-4’s performance was not much better than GPT-3.5. Although GPT-4 may be more accurate, it could have lower recall compared to GPT-3.5 (**Table 3**), and recall is much more important than precision when screening literature for a meta-analysis. Furthermore, when evaluating the performance of three prompt strategies, GPT-4 and GPT-3.5 shared similar performance across all measures, except workload reduction, where GPT-4 showed a slightly better performance (GPT-3.5 vs. GPT-4, medium: 0.303 vs. 0.345; ANOVA test, P = 0.037; **Supplementary Fig 6H**). In short, in the context of this research, neither GPT-3.5 nor GPT-4 was overwhelmingly superior to the other one.

It is important to evaluate the performance of LARS-GPT in various scenarios. Thus, in the study, we selected 4 meta-analyses with distinct type of diseases, which stand for cancer, immune-related disease, metabolic-related disease, and skeletal muscle disorder, respectively. In general, LARS-GPT demonstrated an ideal performance on all of them (Table 4). What really impact the performance of LARS-GPT is the prompts designed, which also highlights the value of prompt design steps in our pipeline.

In this study, a single-prompt is developed from a single filtering criteria, and a key step of single-prompt creation is the selection of criteria. Potential criteria should be derived from the inclusion and exclusion criteria of the designed meta-analysis. In some cases, however, researchers need to extract information from a subgroup analysis, which may not be presented in the title and abstract of a record, *e*.*g*. materials used in surgery,^18^ and criteria related to such information are not suitable for prompt creation. To avoid this issue, it is better to use options that are more likely to be adequately judged using only the title and abstract of record, which are criteria related to “Species”, “Disease”, and “Research type”. In fact, the majority of the best combinations identified in the current research were based on these three criteria. Thus, users are recommended to try them first when using LARS-GPT.

To apply LARS-GPT, users need to manually label a few records for single-prompts so that the best combination can be identified. Based on our experiences, to be well evaluated, each single-prompt needs around 10 positive and 10 negative records. Considering overlaps between the records for single-prompts, researchers need to label about 20 to 100 records for five single-prompts. Once an application based on LARS-GPT is developed, it will be much easier to do this labeling.

We tried three prompt strategies, including a “chain of thought prompt” (prompt strategy 2) that was designed following the OpenAI’s guidelines. Surprisingly, all three prompt strategies showed comparable performance (**Table 4; Supplementary Fig 6A-D**). Indeed, the “chain of thought prompt” takes more time for ChatGPT to answer in a more organized format. However, this improvement does not translate into enhanced performance in LARS-GPT. A possible reason is that the two other “less-structured” strategies already guided ChatGPT sufficiently. However, due to the “black box” nature of ChatGPT, we cannot explain the phenomenon. As a result, users are recommended to select whichever they prefer.

In our research, we did not use metrics like Work Saved over Sampling (WSS) and Average Time to Discover (ATD),^5^ which have been commonly used to evaluate previous AI models. This is because LARS-GPT works in a completely different way, as mentioned before. Within LARS-GPT, ChatGPT will directly answer whether to include or exclude a record, instead of returning a probability for it.

## Conclusion

This study developed a pipeline named LARS-GPT, and using this pipeline showed that an automatic selection of records for a meta-analysis is possible with ChatGPT.

## Supporting information

https://github.com/xiangmingcai/LARS

## Data Availability

The original code used in this paper are available in Github (https://github.com/xiangmingcai/LARS). All responses from ChatGPT can be found in Supplementary File 2. Any additional information required is available from the corresponding author upon request.

## Acknowledgments

We would like to also thank OpenAI for sharing ChatGPT with the research field. This study was funded by the China Scholarship Council (CSC; grant no. 202206090022).

## Author Contributions

XC and CM conceived the idea. XC designed the study. XC, YG, and YD developed the methodology, acquired, and analyzed the data. XC, BW, DW, and JJGV were involved in interpretation of data. XC drafted the manuscript and all authors edited the manuscript. All authors had full access to the raw data in the study. XC and YG accessed and verified the data. XC and JJGV oversaw conduction of the study. All authors contributed to the article and approved the submitted version. All authors had final responsibility for the decision to submit for publication.

## Data sharing

The original code used in this paper are available in Github (https://github.com/xiangmingcai/LARS). All responses from ChatGPT can be found in **Supplementary File 2**. Any additional information required is available from the corresponding author upon request.

## Supplementary materials

**Supplementary Figure 1** The responses of GPT-3.5 show high robustness in classifying records

(**A**) The bar plot shows the robustness score of each single-prompt from the four meta-analyses included. (**B**) The stack bar plot shows the answers of repeated requests sent to GPT-3.5 with single-prompts from glioma meta-analysis. N, no; Y, yes; NS, not sure.

**Supplementary Figure 2** The other returns for robustness evaluation

The stack bar plots show the answers of repeated requests sent to GPT-3.5 with single-prompts from inflammatory bowel diseases (**A**), diabetes mellitus (**B**), and sarcopenia (**C**) meta-analyses, respectively. N, no; Y, yes; NS, not sure.

**Supplementary Figure 3** Performance of all combinations of single-prompts from inflammatory bowel diseases meta-analysis

Precision (**A, E**), recall (**B, F**), F1 score (**C, G**), and workload reduction (**D, H**) are presented for GPT-3.5 (**A-D**) and GPT-4 (**E-F**), respectively. Best combination is marked with a triangle.

**Supplementary Figure 4** Performance of all combinations of single-prompts from diabetes mellitus meta-analysis

Precision (**A, E**), recall (**B, F**), F1 score (**C, G**), and workload reduction (**D, H**) are presented for GPT-3.5 (**A-D**) and GPT-4 (**E-F**), respectively. Best combination is marked with a red triangle. Sub-best combination is marked with a yellow triangle.

**Supplementary Figure 5** Performance of all combinations of single-prompts from sarcopenia meta-analysis

Precision (**A, E**), recall (**B, F**), F1 score (**C, G**), and workload reduction (**D, H**) are presented for GPT-3.5 (**A-D**) and GPT-4 (**E-F**), respectively. Best combination is marked with a triangle.

**Supplementary Figure 6** Comparison of the performance of best combinations

Comparison of the performance between three prompt strategies, regarding precision (**A**), recall (**B**), F1 score (**C**), and workload reduction (**D**). Comparison of the performance between GPT-3.5 and GPT-4, regarding precision (**E**), recall (**F**), F1 score (**G**), and workload reduction (**H**).

**Supplementary Table 1** Performance of single-prompts from inflammatory bowel diseases, diabetes mellitus, and sarcopenia meta-analyses using GPT-3.5, GPT-4, and random classifier

**Supplementary File 1** The content of three prompt strategies using full combination

**Supplementary File 2** Validation datasets from four meta-analyses

**Supplementary File 3** Supplementary Methods

## Reference

1. Subbiah V. The next generation of evidence-based medicine. Nat Med 2023;29(1):49–58. (In eng). DOI: 10.1038/s41591-022-02160-z.

2. Abdelkader W, Navarro T, Parrish R, et al. A Deep Learning Approach to Refine the Identification of High-Quality Clinical Research Articles From the Biomedical Literature: Protocol for Algorithm Development and Validation. JMIR Res Protoc 2021;10(11):e29398. (In eng). DOI: 10.2196/29398.

3. Tercero-Hidalgo JR, Khan KS, Bueno-Cavanillas A, et al. Artificial intelligence in COVID-19 evidence syntheses was underutilized, but impactful: a methodological study. J Clin Epidemiol 2022;148:124–134. (In eng). DOI: 10.1016/j.jclinepi.2022.04.027.

4. Gates A, Gates M, Sebastianski M, Guitard S, Elliott SA, Hartling L. The semi-automation of title and abstract screening: a retrospective exploration of ways to leverage Abstrackr’s relevance predictions in systematic and rapid reviews. BMC Med Res Methodol 2020;20(1):139. (In eng). DOI: 10.1186/s12874-020-01031-w.

5. Ferdinands G, Schram R, de Bruin J, et al. Performance of active learning models for screening prioritization in systematic reviews: a simulation study into the Average Time to Discover relevant records. Syst Rev 2023;12(1):100. (In eng). DOI: 10.1186/s13643-023-02257-7.

6. van Dis EAM, Bollen J, Zuidema W, van Rooij R, Bockting CL. ChatGPT: five priorities for research. Nature 2023;614(7947):224–226. (In eng). DOI: 10.1038/d41586-023-00288-7.

7. Li H, Moon JT, Purkayastha S, Celi LA, Trivedi H, Gichoya JW. Ethics of large language models in medicine and medical research. Lancet Digit Health 2023;5(6):e333–e335. (In eng). DOI: 10.1016/s2589-7500(23)00083-3.

8. Ali SR, Dobbs TD, Hutchings HA, Whitaker IS. Using ChatGPT to write patient clinic letters. Lancet Digit Health 2023;5(4):e179–e181. (In eng). DOI: 10.1016/s2589-7500(23)00048-1.

9. Shaib C, Li M, Joseph S, Marshall IJ, Li JJ, Wallace B. Summarizing, Simplifying, and Synthesizing Medical Evidence Using GPT-3 (with Varying Success). ArXiv 2023;abs/2305.06299.

10. Wang S, Scells H, Koopman B, Zuccon G. Can ChatGPT Write a Good Boolean Query for Systematic Review Literature Search? ArXiv 2023;abs/2302.03495.

11. Borah R, Brown AW, Capers PL, Kaiser KA. Analysis of the time and workers needed to conduct systematic reviews of medical interventions using data from the PROSPERO registry. BMJ Open 2017;7(2):e012545. (In eng). DOI: 10.1136/bmjopen-2016-012545.

12. Talebi S, Zeraattalab-Motlagh S, Rahimlou M, et al. The Association between Total Protein, Animal Protein, and Animal Protein Sources with Risk of Inflammatory Bowel Diseases: A Systematic Review and Meta-Analysis of Cohort Studies. Adv Nutr 2023;14(4):752–761. (In eng). DOI: 10.1016/j.advnut.2023.05.008.

13. Aune D, Schlesinger S, Mahamat-Saleh Y, Zheng B, Udeh-Momoh CT, Middleton LT. Diabetes mellitus, prediabetes and the risk of Parkinson’s disease: a systematic review and meta-analysis of 15 cohort studies with 29.9 million participants and 86,345 cases. Eur J Epidemiol 2023;38(6):591–604. (In eng). DOI: 10.1007/s10654-023-00970-0.

14. Beaudart C, Demonceau C, Reginster JY, et al. Sarcopenia and health-related quality of life: A systematic review and meta-analysis. J Cachexia Sarcopenia Muscle 2023;14(3):1228–1243. (In eng). DOI: 10.1002/jcsm.13243.

15. Page MJ, McKenzie JE, Bossuyt PM, et al. The PRISMA 2020 statement: an updated guideline for reporting systematic reviews. Bmj 2021;372:n71. (In eng). DOI: 10.1136/bmj.n71.

16. Jin Q, Leaman R, Lu Z. Retrieve, Summarize, and Verify: How Will ChatGPT Affect Information Seeking from the Medical Literature? J Am Soc Nephrol 2023 (In eng). DOI: 10.1681/asn.0000000000000166.

17. Stokel-Walker C, Van Noorden R. What ChatGPT and generative AI mean for science. Nature 2023;614(7947):214–216. (In eng). DOI: 10.1038/d41586-023-00340-6.

18. Cai X, Yang J, Zhu J, et al. Reconstruction strategies for intraoperative CSF leak in endoscopic endonasal skull base surgery: systematic review and meta-analysis. Br J Neurosurg 2022;36(4):436–446. (In eng). DOI: 10.1080/02688697.2020.1849548.

