## Supplementary material for "Utilizing ChatGPT to select literature for meta-analysis shows workload reduction while maintaining a similar recall level as manual curation": https://github.com/xiangmingcai/LARS: Supplementary Materials.pdf

**Supplementary Figures** 2

**Supplementary Figure 1** The responses of GPT-3.5 show high robustness in classifying records 2

**Supplementary Figure 2** The other returns for robustness evaluation 3

**Supplementary Figure 3** Performance of all combinations of single-prompts from inflammatory bowel diseases meta-analysis 4

**Supplementary Figure 4** Performance of all combinations of single-prompts from diabetes mellitus meta-analysis 5

**Supplementary Figure 5** Performance of all combinations of single-prompts from sarcopenia meta-analysis 6

**Supplementary Figure 6** Comparison of the performance of best combinations 7

**Supplementary Tables** 8

**Supplementary Table 1** Performance of single-prompts from inflammatory bowel diseases, diabetes mellitus, and sarcopenia meta-analyses using GPT-3.5, GPT-4, and random classifier 8

**Supplementary Files** 10

**Supplementary File 1** The content of three prompt strategies using full combination 10

**Supplementary File 2** Validation datasets from four meta-analyses 18

**Supplementary File 4** Supplementary Methods 19

### Supplementary Figures

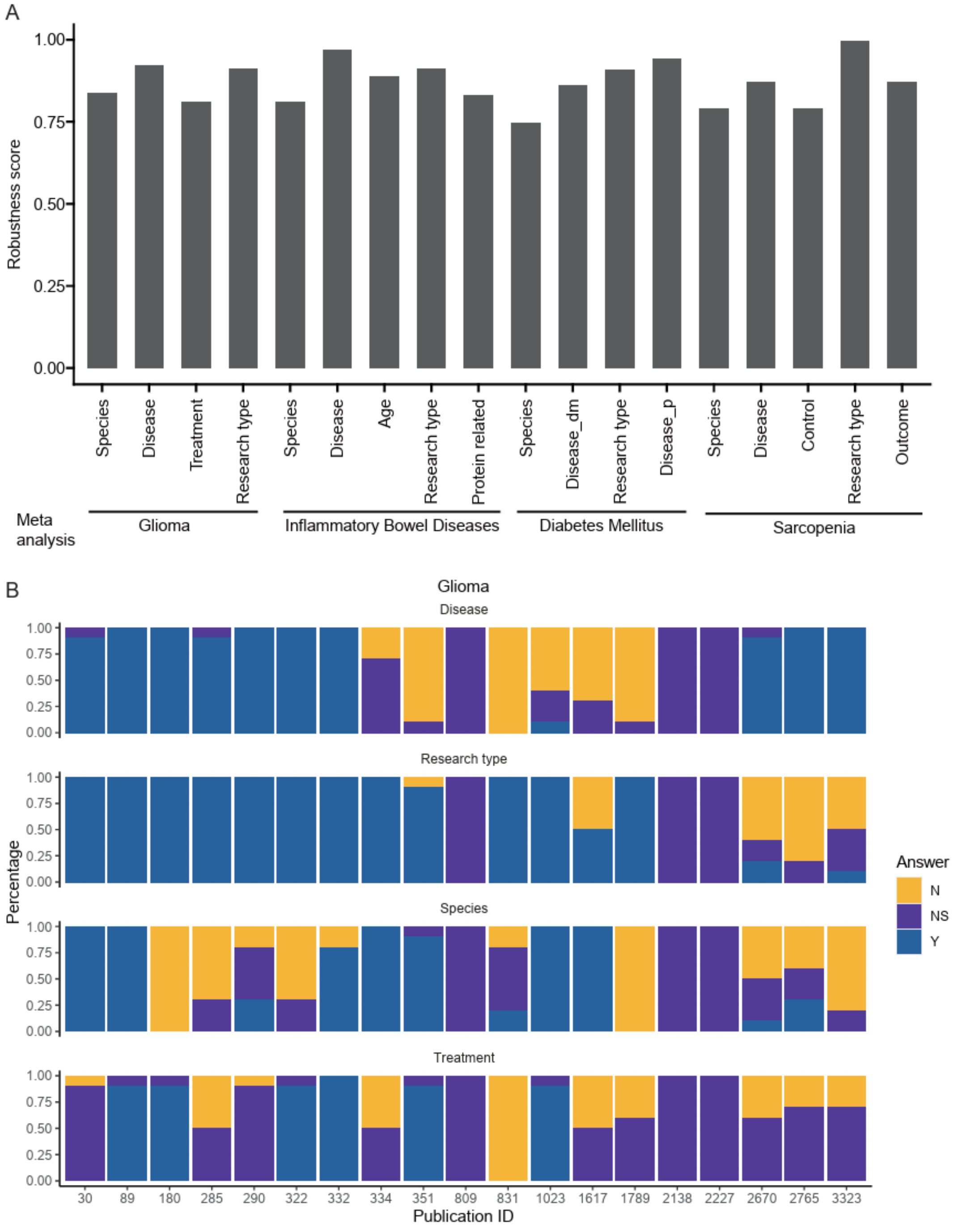


#### Supplementary Figure 1 The responses of GPT-3.5 show high robustness in classifying records

(**A**) The bar plot shows the robustness score of each single-prompt from the four meta-analyses included. (**B**) The stack bar plot shows the answers of repeated requests sent to GPT-3.5 with single-prompts from glioma meta-analysis. N, no; Y, yes; NS, not sure.

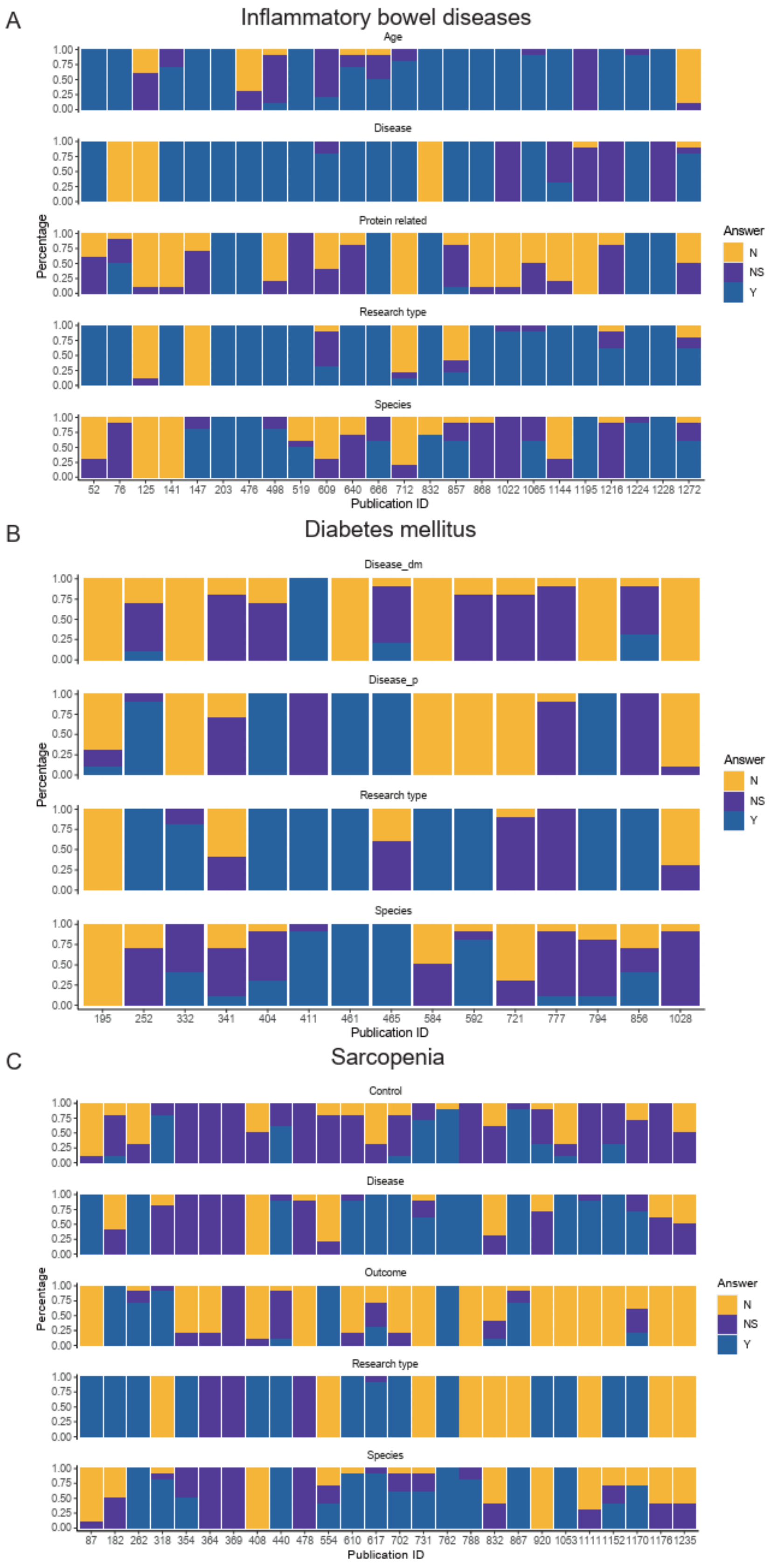


**Supplementary Figure 2** The other returns for robustness evaluation

The stack bar plots show the answers of repeated requests sent to GPT-3.5 with single-prompts from inflammatory bowel diseases (**A**), diabetes mellitus (**B**), and sarcopenia (**C**) meta-analyses, respectively. N, no; Y, yes; NS, not sure.

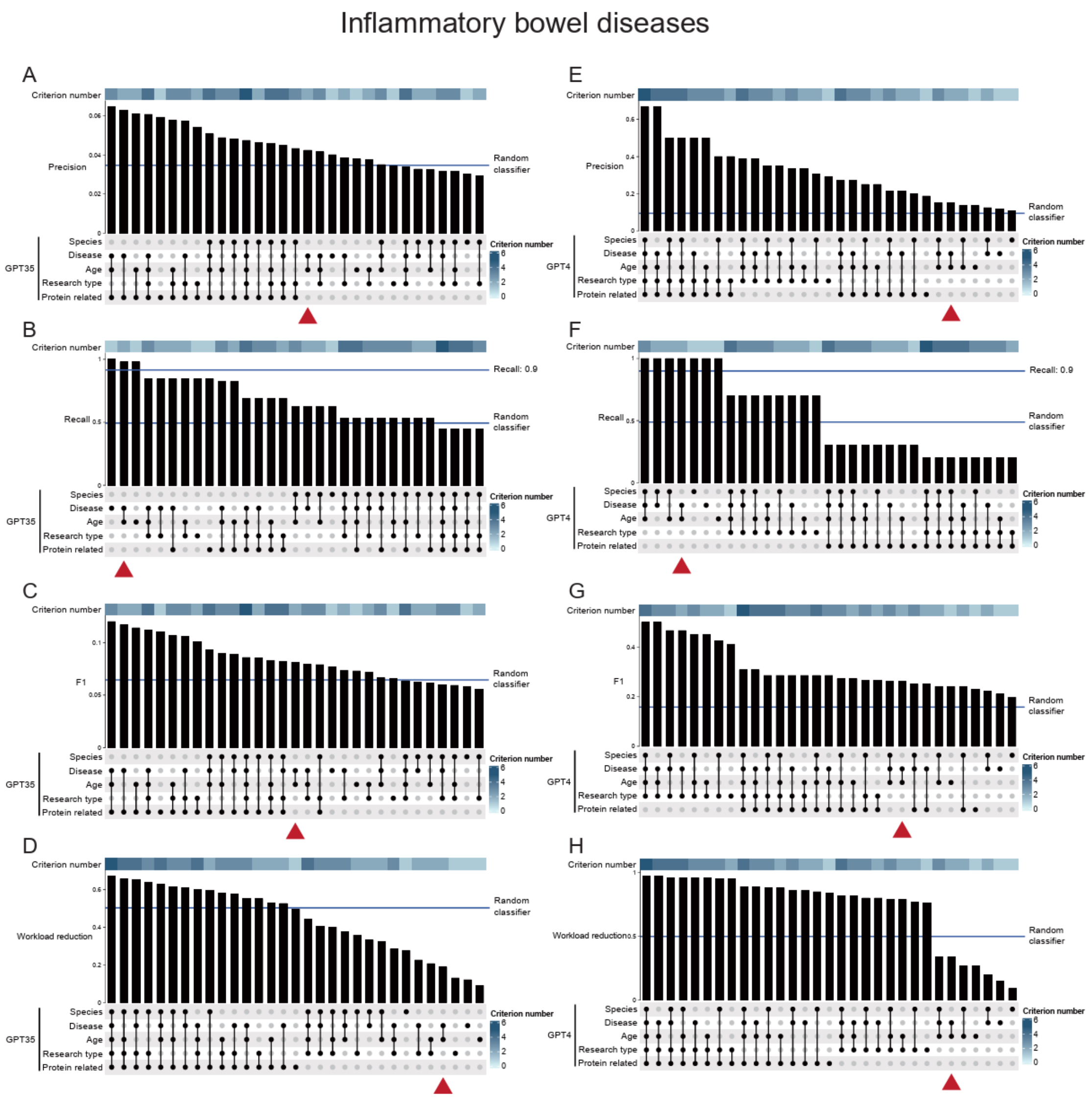


**Supplementary Figure 3** Performance of all combinations of single-prompts from inflammatory bowel diseases meta-analysis

Precision (**A**, **E**), recall (**B**, **F**), F1 score (**C**, **G**), and workload reduction (**D**, **H**) are presented for GPT-3.5 (**A-D**) and GPT-4 (**E-F**), respectively. Best combination is marked with a triangle.

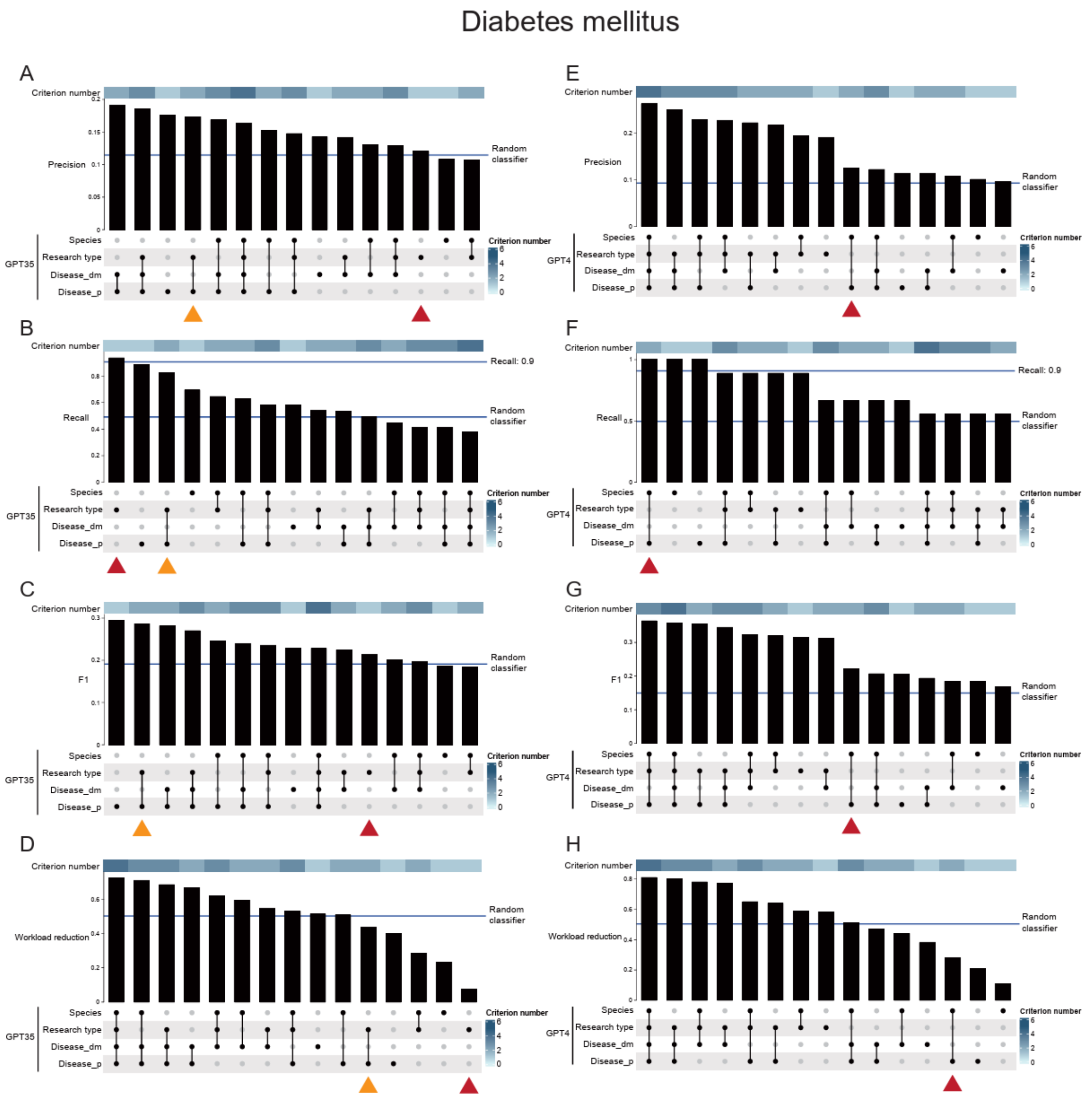


**Supplementary Figure 4** Performance of all combinations of single-prompts from diabetes mellitus meta-analysis

Precision (**A**, **E**), recall (**B**, **F**), F1 score (**C**, **G**), and workload reduction (**D**, **H**) are presented for GPT-3.5 (**A-D**) and GPT-4 (**E-F**), respectively. Best combination is marked with a red triangle. Sub-best combination is marked with a yellow triangle.

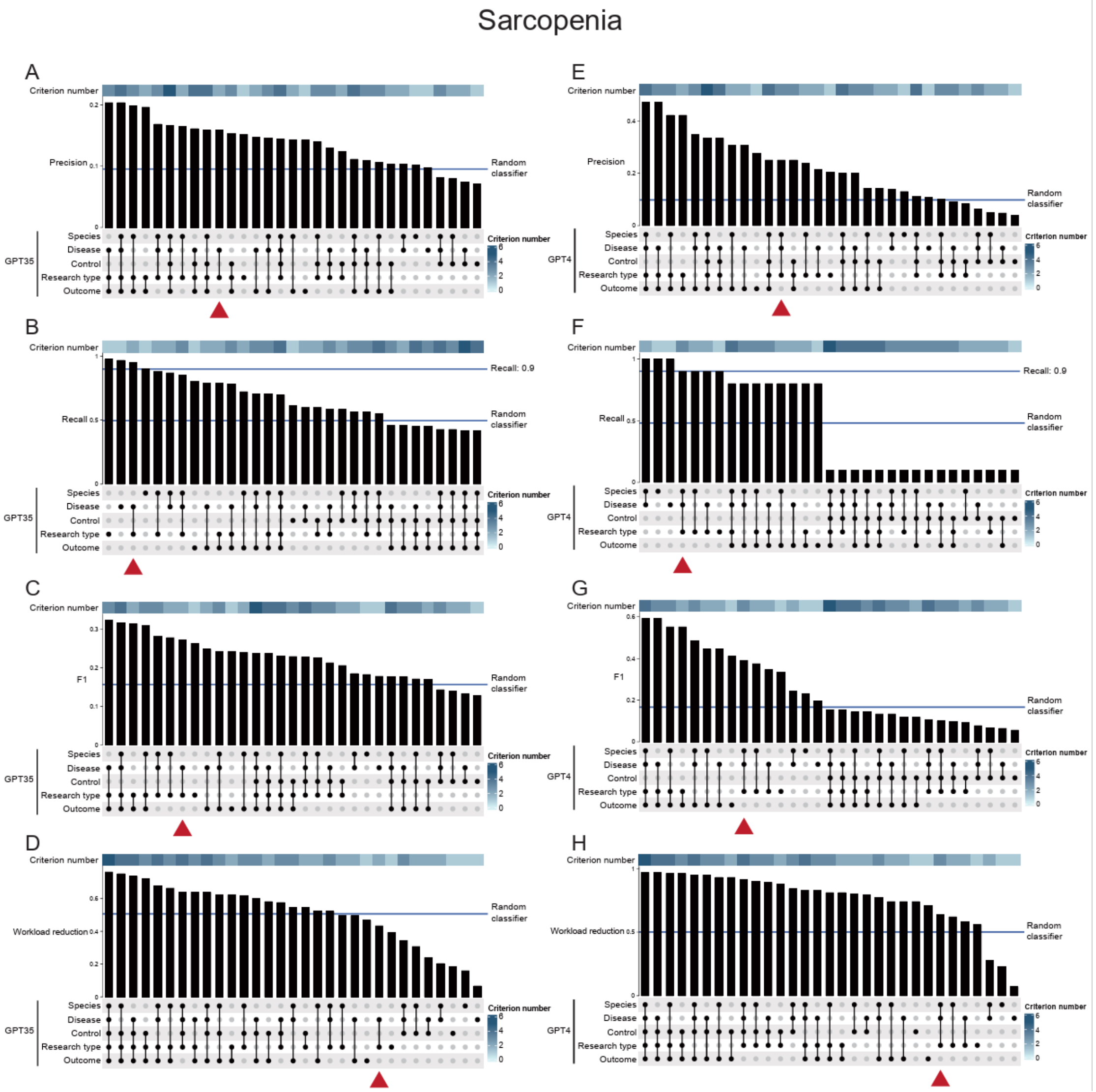


**Supplementary Figure 5** Performance of all combinations of single-prompts from sarcopenia meta-analysis

Precision (**A**, **E**), recall (**B**, **F**), F1 score (**C**, **G**), and workload reduction (**D**, **H**) are presented for GPT-3.5 (**A-D**) and GPT-4 (**E-F**), respectively. Best combination is marked with a triangle.

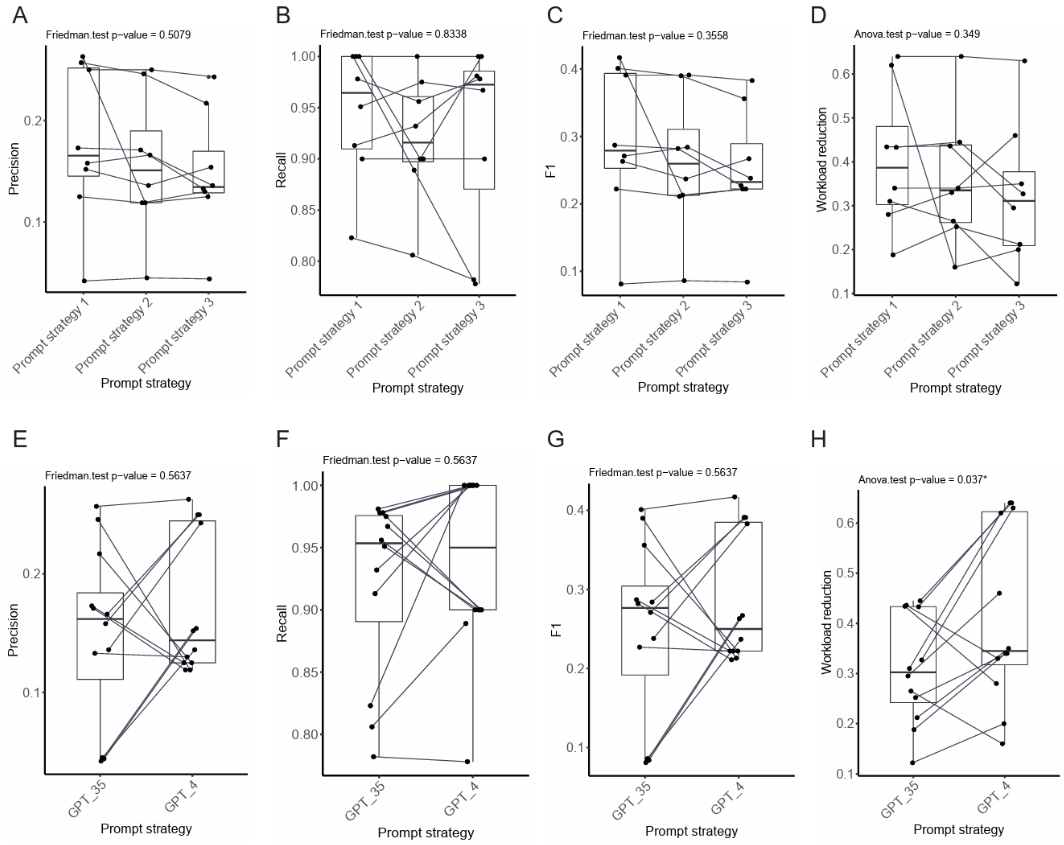


#### Supplementary Figure 6 Comparison of the performance of best combinations

Comparison of the performance between three prompt strategies, regarding precision (**A**), recall (**B**), F1 score (**C**), and workload reduction (**D**). Comparison of the performance between GPT-3.5 and GPT-4, regarding precision (**E**), recall (**F**), F1 score (**G**), and workload reduction (**H**).

### Supplementary Tables

#### **Supplementary Table 1** Performance of single-prompts from inflammatory bowel diseases, diabetes mellitus, and sarcopenia meta-analyses using GPT-3.5, GPT-4, and random classifier

**Inflammatory Bowel Diseases**

| single-prompt | Model | Precision | Recall | F1 | Workload reduction |
|---|---|---|---|---|---|
| Species | GPT-35 | 0.973 | 0.780 | 0.866 | 0.270 |
| | GPT-4 | 0.989 | 0.989 | 0.989 | 0.090 |
| | Random classifier | 0.909 | 0.496 | 0.641 | 0.504 |
| Disease | GPT-35 | 0.911 | 0.965 | 0.937 | 0.100 |
| | GPT-4 | 0.965 | 0.965 | 0.965 | 0.150 |
| | Random classifier | 0.856 | 0.509 | 0.637 | 0.494 |
| Research type | GPT-35 | 0.241 | 1.000 | 0.389 | 0.130 |
| | GPT-4 | 0.750 | 0.857 | 0.800 | 0.760 |
| | Random classifier | 0.201 | 0.479 | 0.283 | 0.501 |
| Age | GPT-35 | 0.819 | 1.000 | 0.901 | 0.060 |
| | GPT-4 | 1.000 | 0.948 | 0.973 | 0.270 |
| | Random classifier | 0.775 | 0.497 | 0.604 | 0.506 |
| Protein related | GPT-35 | 0.500 | 0.897 | 0.642 | 0.480 |
| | GPT-4 | 0.875 | 0.483 | 0.622 | 0.840 |
| | Random classifier | 0.287 | 0.482 | 0.359 | 0.513 |

**Diabetes Mellitus**

| single-prompt | Model | Precision | Recall | F1 | Workload reduction |
|---|---|---|---|---|---|
| Species | GPT-35 | 0.975 | 0.823 | 0.893 | 0.190 |
| | GPT-4 | 0.989 | 0.917 | 0.951 | 0.110 |
| | Random classifier | 0.957 | 0.494 | 0.651 | 0.504 |
| Research type | GPT-35 | 0.449 | 1.000 | 0.620 | 0.110 |
| | GPT-4 | 0.810 | 0.850 | 0.829 | 0.580 |
| | Random classifier | 0.393 | 0.497 | 0.438 | 0.494 |
| Disease_dm | GPT-35 | 0.837 | 0.554 | 0.667 | 0.510 |
| | GPT-4 | 0.919 | 0.770 | 0.838 | 0.380 |
| | Random classifier | 0.739 | 0.499 | 0.594 | 0.501 |
| Disease_p | GPT-35 | 0.930 | 0.663 | 0.774 | 0.430 |
| | GPT-4 | 0.937 | 0.925 | 0.931 | 0.210 |
| | Random classifier | 0.801 | 0.494 | 0.609 | 0.506 |

**Sarcopenia**

| single-prompt | Model | Precision | Recall | F1 | Workload reduction |
|---|---|---|---|---|---|
| Species | GPT-35 | 0.939 | 0.837 | 0.885 | 0.180 |
| | GPT-4 | 0.987 | 0.826 | 0.899 | 0.230 |
| | Random classifier | 0.920 | 0.496 | 0.643 | 0.504 |
| Disease | GPT-35 | 0.733 | 0.904 | 0.810 | 0.100 |
| | GPT-4 | 0.742 | 0.945 | 0.831 | 0.070 |
| | Random classifier | 0.734 | 0.509 | 0.600 | 0.494 |
| Control | GPT-35 | 0.704 | 0.838 | 0.765 | 0.190 |
| | GPT-4 | 0.615 | 0.235 | 0.340 | 0.740 |
| | Random classifier | 0.678 | 0.497 | 0.572 | 0.501 |
| Research type | GPT-35 | 0.852 | 1.000 | 0.920 | 0.390 |
| | GPT-4 | 0.977 | 0.827 | 0.896 | 0.560 |
| | Random classifier | 0.517 | 0.492 | 0.502 | 0.506 |

| | | | | | |
|---|---|---|---|---|---|
| Outcome | GPT-35 | 0.490 | 0.774 | 0.600 | 0.510 |
| | GPT-4 | 0.793 | 0.742 | 0.767 | 0.710 |
| | Random classifier | 0.317 | 0.498 | 0.386 | 0.513 |

### Supplementary Files

#### **Supplementary File 1** The content of three prompt strategies using full combination

**Meta analysis: Glioma**

**Prompt with single criterion (single-prompt) / Prompt strategy 1 (Full combination)**

**Species**

I want you to act as a helpful assistant. I will give you title and abstract of a publication and you will reply whether it meets our criteria or not. I want you to only reply with yes, no, or not sure, and followed with reasons. The criteria is: studies that use human as primary research subject.

**Disease**

I want you to act as a helpful assistant. I will give you title and abstract of a publication and you will reply whether it meets our criteria or not. I want you to only reply with yes, no, or not sure, and followed with reasons. The criteria is: studies that involve patients with glioma, glioblastoma, astrocytoma, oligodendroglioma.

**Treatment**

I want you to act as a helpful assistant. I will give you title and abstract of a publication and you will reply whether it meets our criteria or not. I want you to only reply with yes, no, or not sure, and followed with reasons. The criteria is: The title and abstract must explicitly mention that the study involves treatment of patients with one or more of the following immunotherapies: Immune Checkpoint Inhibitors (e.g. anti-PD-1, anti-PD-L1, anti-CTLA-4), Peptide Vaccination, Dendritic cell Vaccination, Adoptive Transfer of Effector Lymphocytes (e.g. CAR-T cell therapy, TCR-engineered T cells), Chimeric Antigen Receptor (CAR) T-cell therapy, or Oncolytic Viral Vectors therapy.

**Research type**

I want you to act as a helpful assistant. I will give you title and abstract of a publication and you will reply whether it meets our criteria or not. I want you to only reply with yes, no, or not sure, and followed with reasons. The criteria is: studies that was original research instead of reviews, protocol, or case report with less than five patients.

**Chain of thought prompt (Prompt strategy 2; Full combination)**

I want you to act as a helpful assistant. I will give you title and abstract of a publication and I want you to perform the following actions:

1. you will judge whether the publication meets the first criteria or not. The first criteria is: studies that use human as primary research subject.

2. you will judge whether the publication meets the second criteria or not. The second criteria is: studies that involve patients with glioma, glioblastoma, astrocytoma, oligodendroglioma.

3. you will judge whether the publication meets the third criteria or not. The third criteria is: The title and abstract must explicitly mention that the study involves treatment of patients with one or more of the following immunotherapies: Immune Checkpoint Inhibitors (e.g. anti-PD-1, anti-PD-L1, anti-CTLA-4), Peptide Vaccination, Dendritic cell Vaccination, Adoptive Transfer of Effector Lymphocytes (e.g. CAR-T cell therapy, TCR-engineered T cells), Chimeric Antigen Receptor (CAR) T-cell therapy, or Oncolytic Viral Vectors therapy.

4. you will judge whether the publication meets the fourth criteria or not. The fourth criteria is: studies that was original research instead of reviews, protocol, or case report with less than five patients.

5. you will summary the judgements from action 1 to 4. If the publication doesn't meet one or more of these criteria, you will answer 'No', otherwise, you will answer 'Yes'. Note that 'unclear' is treated as 'Yes'.

6.output a json object that contains the following keys: 'answer of action 1', 'answer of action 2', 'answer of action 3', 'answer of action 4', 'answer of action 5'.

**Instruction prompt (Prompt strategy 3; Full combination)**

I want you to act as a helpful assistant. I will give you title and abstract of a publication and you will reply whether it meets all these following criteria or not: (1) studies that use human as primary research subject; (2) studies that involve patients with glioma, glioblastoma, astrocytoma, oligodendroglioma; (3) The title and abstract must explicitly mention that the study involves treatment of patients with one or more of the following immunotherapies: Immune Checkpoint Inhibitors (e.g. anti-PD-1, anti-PD-L1, anti-CTLA-4), Peptide Vaccination, Dendritic cell Vaccination, Adoptive Transfer of Effector Lymphocytes (e.g. CAR-T cell therapy, TCR-engineered T cells), Chimeric Antigen Receptor (CAR) T-cell therapy, or Oncolytic Viral Vectors therapy; (4) studies that was original research instead of reviews, protocol, or case report with less than five patients. Please answer in this way: If the publication doesn't meet one or more of these criteria, you will answer 'No', otherwise, you will answer 'Yes'. Note that 'unclear' is treated as 'Yes'.

**Meta analysis: Inflammatory Bowel Diseases**

**Prompt with single criterion (single-prompt) / Prompt strategy 1 (Full combination)**

**Species**

I want you to act as a helpful assistant. I will give you title and abstract of a publication and you will reply whether it meets our criteria or not. I want you to only reply with yes, no, or not sure, and followed with reasons. The criteria is: studies that only use human as research subject.

**Disease**

I want you to act as a helpful assistant. I will give you title and abstract of a publication and you will reply whether it meets our criteria or not. I want you to only reply with yes, no, or not sure, and followed with reasons. The criteria is: studies that involve patients with inflammatory bowel diseases, including Crohn's disease and ulcerative colitis.

**Research type**

I want you to act as a helpful assistant. I will give you title and abstract of a publication and you will reply whether it meets our criteria or not. I want you to only reply with yes, no, or not sure, and followed with reasons. The criteria is: studies that are prospective cohort study instead of case-contral study, cross-sectional study, retrospective cohort study, randomized controlled trial, review, or protocol.

**Age**

I want you to act as a helpful assistant. I will give you title and abstract of a publication and you will reply whether it meets our criteria or not. I want you to only reply with yes, no, or not sure, and followed with reasons. The criteria is: studies that involve adult patients (at least 18 years old).

**Protein related**

I want you to act as a helpful assistant. I will give you title and abstract of a publication and you will reply whether it meets our criteria or not. I want you to only reply with yes, no, or not sure, and followed with reasons. The criteria is: The title and abstract must mention that the study is related to the consumption of protein (e.g., total dairy, milk, meat, fish, poultry, process meat, and egg).

**Chain of thought prompt (Prompt strategy 2; Full combination)**

I want you to act as a helpful assistant. I will give you title and abstract of a publication and I want you to perform the following actions:

1. you will judge whether the publication meets the first criteria or not. The first criteria is: studies that only use human as research subject.

2. you will judge whether the publication meets the second criteria or not. The second criteria is: studies that involve patients with inflammatory bowel diseases, including Crohn's disease and ulcerative colitis.

3. you will judge whether the publication meets the third criteria or not. The third criteria is: studies that are prospective cohort study instead of case-contral study, cross-sectional study, retrospective cohort study, randomized controlled trial, review, or protocol.

4. you will judge whether the publication meets the fourth criteria or not. The fourth criteria is: studies that involve adult patients (at least 18 years old).

5. you will judge whether the publication meets the fifth criteria or not. The fifth criteria is: The title and abstract must mention that the study is related to the consumption of protein (e.g., total dairy, milk, meat, fish, poultry, process meat, and egg).

6. you will summary the judgements from action 1 to 5. If the publication doesn't meet one or more of these criteria, you will answer 'No', otherwise, you will answer 'Yes'. Note that 'unclear' is treated as 'Yes'.

7.output a json object that contains the following keys: 'answer of action 1', 'answer of action 2', 'answer of action 3', 'answer of action 4', 'answer of action 5', 'answer of action 6'.

**Instruction prompt (Prompt strategy 3; Full combination)**

I want you to act as a helpful assistant. I will give you title and abstract of a publication and you will reply whether it meets all these following criteria or not: (1) studies that only use human as research subject; (2) studies that involve patients with inflammatory bowel diseases, including Crohn's disease and ulcerative colitis; (3) studies that are prospective cohort study instead of case-contral study, cross-sectional study, retrospective cohort study, randomized controlled trial, review, or protocol; (4) studies that involve adult patients (at least 18 years old); (5) The title and abstract must mention that the study is related to the consumption of protein (e.g., total dairy, milk, meat, fish, poultry, process meat, and egg). Please answer in this way: If the publication doesn't meet one or more of these criteria, you will answer 'No', otherwise, you will answer 'Yes'. Note that 'unclear' is treated as 'Yes'.

**Meta analysis: Diabetes Mellitus**

**Prompt with single criterion (single-prompt) / Prompt strategy 1 (Full combination)**

**Species**

I want you to act as a helpful assistant. I will give you title and abstract of a publication and you will reply whether it meets our criteria or not. I want you to only reply with yes, no, or not sure, and followed with reasons. The criteria is: studies that only use human as research subject.

**Research type**

I want you to act as a helpful assistant. I will give you title and abstract of a publication and you will reply whether it meets our criteria or not. I want you to only reply with yes, no, or not sure, and followed with reasons. The criteria is: studies that are prospective or retrospective cohort study, case-control study. Of note, these research types doesn't meet the criteria: cross-sectional study, randomized controlled trial, review, protocol or others.

**Disease_dm**

I want you to act as a helpful assistant. I will give you title and abstract of a publication and you will reply whether it meets our criteria or not. I want you to only reply with yes, no, or not sure, and followed with reasons. The criteria is: studies that involve patients with diabetes mellitus.

**Disease_p**

I want you to act as a helpful assistant. I will give you title and abstract of a publication and you will reply whether it meets our criteria or not. I want you to only reply with yes, no, or not sure, and followed with reasons. The criteria is: studies that involve patients with parkinson's disease.

**Chain of thought prompt (Prompt strategy 2; Full combination)**

I want you to act as a helpful assistant. I will give you title and abstract of a publication and I want you to perform the following actions:

1. you will judge whether the publication meets the first criteria or not. The first criteria is: studies that only use human as research subject.

2. you will judge whether the publication meets the second criteria or not. The second criteria is: studies that are prospective or retrospective cohort study, case-control study. Of note, these research types doesn't meet the criteria: cross-sectional study, randomized controlled trial, review, protocol or others.

3. you will judge whether the publication meets the third criteria or not. The third criteria is: studies that involve patients with diabetes mellitus.

4. you will judge whether the publication meets the fourth criteria or not. The fourth criteria is: studies that involve patients with parkinson's disease.

5. you will summary the judgements from action 1 to 4. If the publication doesn't meet one or more of these criteria, you will answer 'No', otherwise, you will answer 'Yes'. Note that 'unclear' is treated as 'Yes'.

6.output a json object that contains the following keys: 'answer of action 1', 'answer of action 2', 'answer of action 3', 'answer of action 4', 'answer of action 5'.

**Instruction prompt (Prompt strategy 3; Full combination)**

I want you to act as a helpful assistant. I will give you title and abstract of a publication and you will reply whether it meets all these following criteria or not: (1)studies that only use human as research subject; (2) studies that are prospective or retrospective cohort study, case-control study. Of note, these research types doesn't meet the criteria: cross-sectional study, randomized controlled trial, review, protocol or others; (3)studies that involve patients with diabetes mellitus; (4) studies that involve patients with parkinson's disease. Please answer in this way: If the publication doesn't meet one or more of these criteria, you will answer 'No', otherwise, you will answer 'Yes'. Note that 'unclear' is treated as 'Yes'.

**Meta analysis: Sarcopenia**

**Prompt with single criterion (single-prompt) / Prompt strategy 1 (Full combination)**

**Species**

I want you to act as a helpful assistant. I will give you title and abstract of a publication and you will reply whether it meets our criteria or not. I want you to only reply with yes, no, or not sure, and followed with reasons. The criteria is: studies that only use human as research subject.

**Disease**

I want you to act as a helpful assistant. I will give you title and abstract of a publication and you will reply whether it meets our criteria or not. I want you to only reply with yes, no, or not sure, and followed with reasons. The criteria is: studies that involve patients with sarcopenia.

**Control**

I want you to act as a helpful assistant. I will give you title and abstract of a publication and you will reply whether it meets our criteria or not. I want you to only reply with yes, no, or not sure, and followed with reasons. The criteria is: studies that involve participants without sarcopenia.

**Research type**

I want you to act as a helpful assistant. I will give you title and abstract of a publication and you will reply whether it meets our criteria or not. I want you to only reply with yes, no, or not sure, and followed with reasons. The criteria is: studies that was original research instead of reviews, protocol, or others.

**Outcome**

I want you to act as a helpful assistant. I will give you title and abstract of a publication and you will reply whether it meets our criteria or not. I want you to only reply with yes, no, or not sure, and followed with reasons. The criteria is: The title and abstract must explicitly mention that the study reported health-related quality of life assessment.

**Chain of thought prompt (Prompt strategy 2; Full combination)**

I want you to act as a helpful assistant. I will give you title and abstract of a publication and I want you to perform the following actions:

1. you will judge whether the publication meets the first criteria or not. The first criteria is: studies that only use human as research subject.

2. you will judge whether the publication meets the second criteria or not. The second criteria is: studies that involve patients with sarcopenia.

3. you will judge whether the publication meets the third criteria or not. The third criteria is: studies that involve participants without sarcopenia.

4. you will judge whether the publication meets the fourth criteria or not. The fourth criteria is: studies that was original research instead of reviews, protocol, or others.

5. you will judge whether the publication meets the fifth criteria or not. The fifth criteria is: The title and abstract must explicitly mention that the study reported health-related quality of life assessment.

6. you will summary the judgements from action 1 to 5. If the publication doesn't meet one or more of these criteria, you will answer 'No', otherwise, you will answer 'Yes'. Note that 'unclear' is treated as 'Yes'.

7.output a json object that contains the following keys: 'answer of action 1', 'answer of action 2', 'answer of action 3', 'answer of action 4', 'answer of action 5', 'answer of action 6'.

**Instruction prompt (Prompt strategy 3; Full combination)**

I want you to act as a helpful assistant. I will give you title and abstract of a publication and you will reply whether it meets all these following criteria or not: (1) studies that only use human as research subject; (2) studies that involve patients with sarcopenia; (3) studies that involve participants without sarcopenia; (4) studies that was original research instead of reviews, protocol, or others; (5) The title and abstract must explicitly mention that the study reported health-related quality of life assessment. Please answer in this way: If the publication doesn't meet one or more of these criteria, you will answer 'No', otherwise, you will answer 'Yes'. Note that 'unclear' is treated as 'Yes'.

#### **Supplementary File 2** Validation datasets from four meta-analyses

See separate file.

#### Supplementary File 4 Supplementary Methods

#### Supplementary method of LARS pipeline

##### Selection of validation records from the inflammatory bowel diseases (IBD), diabetes mellitus (DM), and sarcopenia meta-analyses

Based on the detailed methods from these 3 published meta-analyses, we successfully repeated their literature research and classification of records. For these 3 validation datasets, all records were used for GPT-3.5 evaluation. However, only 100 randomly selected records were used for the GPT-4 evaluation, due to the limited funding. To do the randomization, each record was assigned a randomized number with the RAND function from EXCEL. The records with the top 100 randomized numbers were selected. The percentage of positive records (definition of positive/negative can be found in the Methods section of the manuscript) was checked after randomization. The percentages of positive records are around 10% for all three validation datasets (9 or 10 out of 100).

##### Selection of validation records from the glioma meta-analysis

Due to the large number of records (6020) identified from PubMed in the glioma meta-analysis, which was too much for evaluating ChatGPT, we opted to select a subset of these records. At first, we randomly selected 100 records for GPT-4 analysis (96 negative and 4 positive records). To make sure the glioma dataset has similar percentage of positive records as other validation datasets (around 10%), we randomly selected another 6 positive records to replace 6 randomly selected negative records in this dataset with 100 records. Additionally, we randomly selected 1000 negative records, together with these previously selected 96 negative records to test GPT-3.5. Also, all 264 positive records, which were identified from PubMed, were also included in the glioma validation dataset for GPT-3.5. In the end, a total of 1360 records were used for GPT-3.5 evaluation (**Table 2**).

##### Evaluation of response robustness of ChatGPT

Because of the nature of LLMs, the generated answer from LLMs varies each time, even with exactly identical input. We assessed the robustness of ChatGPT's answers using single-prompts before assessing the LARS-GPT pipeline. In other words, the robustness evaluation is a pre-assessment for ChatGPT. It's not a part of the LARS-GPT pipeline.

To evaluate the robustness of all three different answers ("Yes", "No", "Not sure"), we applied single-prompts on all records before testing the robustness. For each single-prompt, we randomly selected three records for each type of answer ("Yes", "No", "Not sure). Because of the overlap of randomly selected records, deduplication of selected records is conducted. In the end, a total of 74 records were used. And 10 repeated requests for each record were sent to ChatGPT for robustness assessment (see Fig below).

Then, for each single-prompt, we have:

$$\text{Robustness score} = \frac{\sum_{k=1}^{n} \max\left(P_k(Yes), P_k(No), P_k(Not\ sure)\right)}{n}$$

Where n is the number of selected records and P is the abbreviation of probability.

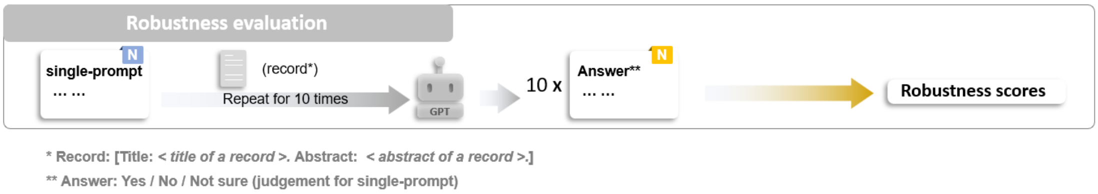


##### Statistical analysis

The random classifier was used as a baseline reference. This classifier randomly assigned an answer of "Yes" or "No" to each record, with 50% chance for each option. To obtain a stable baseline performance, the evaluation for the random classifier was iterated 100 times. The recall and workload reduction of random classifier were both around 0.5, which was set as a reference level.

Python was used to access ChatGPT. All statistical analyses were performed in R software (version 4.1). Upset plot was drawn with

"ComplexHeatmap" R package (version 2.10). Shapiro-Wilk test and Levene's test were applied to check the normality and the homogeneity of variance across groups, respectively. If a normality assumption was met, repeated measures ANOVA test ("rstatix" R package, version 0.7) was used to compare performance between groups. Otherwise, Friedman rank sum test would be used ("stats" R package, version 4.1) for comparison. $P < 0.05$ was applied as indicator for statistical significance.
