## Supplementary material for "Utilizing ChatGPT to select literature for meta-analysis shows workload reduction while maintaining a similar recall level as manual curation": https://github.com/xiangmingcai/LARS: Table 2 meta_summary.pdf

| First author | Field | Publication year | Journal | Original research | | | Our repetition (validation datasets) | |
|---|---|---|---|---|---|---|---|---|
| | | | | All identified records | Identified records from Medline/PubMed | Records preserved in title and abstract screen step | Identified records from Medline/PubMed | Records preserved in title and abstract screen step (Matched) |
| Cai X. | Glioma | Current research | | 8550 | 6020 | 272 | 1360 | 264 |
| Talebi S. | Inflammatory Bowel Diseases | 2023 | Adv Nutr | 2755 | 1285 | 51 | 1284 | 45 |
| Aune D. | Diabetes Mellitus | 2023 | Eur J Epidemiol | 5320 | 1040 | 216 | 1039 | 124 |
| Beaudart C. | Sarcopenia | 2023 | J Cachexia Sarcopenia Muscle | 2293 | NA | 188 | 1293 | 122 |
