## Supplementary material for "Utilizing ChatGPT to select literature for meta-analysis shows workload reduction while maintaining a similar recall level as manual curation": https://github.com/xiangmingcai/LARS: Table 3 prompt with single criterion.docx

| **Glioma** |  |  |  |  |  |
| --- | --- | --- | --- | --- | --- |
| **single-prompt** | **Model** | **Precision** | **Recall** | **F1** | **Workload reduction** |
| Species | GPT-35 | 0.587 | 0.786 | 0.672 | 0.250 |
|  | GPT-4 | 0.791 | 0.607 | 0.687 | 0.570 |
|  | Random classifier | 0.557 | 0.494 | 0.523 | 0.504 |
| Disease | GPT-35 | 0.989 | 0.905 | 0.945 | 0.130 |
|  | GPT-4 | 1.000 | 1.000 | 1.000 | 0.050 |
|  | Random classifier | 0.950 | 0.506 | 0.659 | 0.494 |
| Treatment | GPT-35 | 0.530 | 0.917 | 0.672 | 0.170 |
|  | GPT-4 | 0.745 | 0.792 | 0.768 | 0.490 |
|  | Random classifier | 0.485 | 0.504 | 0.494 | 0.501 |
| Research type | GPT-35 | 0.915 | 0.966 | 0.940 | 0.060 |
|  | GPT-4 | 0.946 | 0.989 | 0.967 | 0.070 |
|  | Random classifier | 0.893 | 0.495 | 0.635 | 0.506 |
