## Supplementary material for "Utilizing ChatGPT to select literature for meta-analysis shows workload reduction while maintaining a similar recall level as manual curation": https://github.com/xiangmingcai/LARS: Table 4 3 prompt strategies.pdf

| Glioma<br>Model | Prompt strategy | Best Combination<br>Precision | Recall | F1 | Workload reduction | Full Combination<br>Precision | Recall | F1 | Workload reduction |
|---|---|---|---|---|---|---|---|---|---|
| GPT-3.5 | Prompt strategy 1 | 0.257 | 0.913 | 0.401 | 0.310 | 0.296 | 0.830 | 0.436 | 0.456 |
| | Prompt strategy 2 | 0.246 | 0.932 | 0.390 | 0.265 | 0.471 | 0.772 | 0.585 | 0.678 |
| | Prompt strategy 3 | 0.217 | 0.981 | 0.356 | 0.122 | 0.358 | 0.852 | 0.504 | 0.538 |
| Random classifier | | 0.194 | 0.499 | 0.280 | 0.502 | 0.194 | 0.499 | 0.280 | 0.502 |
| GPT-4 | Prompt strategy 1 | 0.263 | 1.000 | 0.417 | 0.620 | 0.400 | 0.800 | 0.533 | 0.800 |
| | Prompt strategy 2 | 0.119 | 1.000 | 0.213 | 0.160 | 0.159 | 0.700 | 0.259 | 0.560 |
| | Prompt strategy 3 | 0.125 | 1.000 | 0.222 | 0.200 | 0.233 | 0.700 | 0.350 | 0.700 |
| Random classifier | | 0.100 | 0.495 | 0.166 | 0.504 | 0.100 | 0.495 | 0.166 | 0.504 |

| Inflammatory Bowel Diseases<br>Model | Prompt strategy | Best Combination<br>Precision | Recall | F1 | Workload reduction | Full Combination<br>Precision | Recall | F1 | Workload reduction |
|---|---|---|---|---|---|---|---|---|---|
| GPT-3.5 | Prompt strategy 1 | 0.042 | 0.978 | 0.081 | 0.188 | 0.047 | 0.444 | 0.086 | 0.670 |
| | Prompt strategy 2 | 0.045 | 0.956 | 0.086 | 0.252 | 0.180 | 0.533 | 0.270 | 0.896 |
| | Prompt strategy 3 | 0.044 | 0.978 | 0.084 | 0.212 | 0.084 | 0.778 | 0.152 | 0.675 |
| Random classifier | | 0.036 | 0.514 | 0.067 | 0.502 | 0.036 | 0.514 | 0.067 | 0.502 |
| GPT-4 | Prompt strategy 1 | 0.152 | 1.000 | 0.263 | 0.340 | 0.500 | 0.200 | 0.286 | 0.960 |
| | Prompt strategy 2 | 0.136 | 0.900 | 0.237 | 0.340 | 1.000 | 0.200 | 0.333 | 0.980 |
| | Prompt strategy 3 | 0.154 | 1.000 | 0.267 | 0.350 | 0.500 | 0.100 | 0.167 | 0.980 |
| Random classifier | | 0.098 | 0.485 | 0.163 | 0.504 | 0.098 | 0.485 | 0.163 | 0.504 |

| Diabetes Mellitus<br>Model | Prompt strategy | Sub-best Combination<br>Precision | Recall | F1 | Workload reduction | Full Combination<br>Precision | Recall | F1 | Workload reduction |
|---|---|---|---|---|---|---|---|---|---|
| GPT-3.5 | Prompt strategy 1 | 0.173 | 0.823 | 0.287 | 0.434 | 0.165 | 0.379 | 0.230 | 0.726 |
| | Prompt strategy 2 | 0.171 | 0.806 | 0.282 | 0.436 | 0.520 | 0.106 | 0.176 | 0.976 |
| | Prompt strategy 3 | 0.133 | 0.782 | 0.227 | 0.295 | 0.150 | 0.847 | 0.255 | 0.322 |
| Random classifier | | 0.118 | 0.492 | 0.190 | 0.502 | 0.118 | 0.492 | 0.190 | 0.502 |
| GPT-4 | Prompt strategy 1 | 0.125 | 1.000 | 0.222 | 0.280 | 0.263 | 0.556 | 0.357 | 0.810 |
| | Prompt strategy 2 | 0.119 | 0.889 | 0.211 | 0.330 | 0.222 | 0.444 | 0.296 | 0.820 |
| | Prompt strategy 3 | 0.130 | 0.778 | 0.222 | 0.460 | 0.143 | 0.667 | 0.235 | 0.580 |
| Random classifier | | 0.089 | 0.491 | 0.151 | 0.504 | 0.089 | 0.491 | 0.151 | 0.504 |

| Sarcopenia<br>Model | Prompt strategy | Best Combination<br>Precision | Recall | F1 | Workload reduction | Full Combination<br>Precision | Recall | F1 | Workload reduction |
|---|---|---|---|---|---|---|---|---|---|
| GPT-3.5 | Prompt strategy 1 | 0.158 | 0.951 | 0.271 | 0.433 | 0.166 | 0.418 | 0.237 | 0.762 |
| | Prompt strategy 2 | 0.166 | 0.975 | 0.284 | 0.445 | 0.286 | 0.148 | 0.195 | 0.951 |
| | Prompt strategy 3 | 0.136 | 0.967 | 0.238 | 0.327 | 0.208 | 0.902 | 0.337 | 0.590 |
| Random classifier | | 0.093 | 0.493 | 0.157 | 0.501 | 0.093 | 0.493 | 0.157 | 0.501 |
| GPT-4 | Prompt strategy 1 | 0.250 | 0.900 | 0.391 | 0.640 | 0.333 | 0.100 | 0.154 | 0.970 |
| | Prompt strategy 2 | 0.250 | 0.900 | 0.391 | 0.640 | 0.500 | 0.600 | 0.545 | 0.880 |
| | Prompt strategy 3 | 0.243 | 0.900 | 0.383 | 0.630 | 0.500 | 0.500 | 0.500 | 0.900 |
| Random classifier | | 0.099 | 0.488 | 0.164 | 0.504 | 0.099 | 0.488 | 0.164 | 0.504 |
