## Supplementary figures and images for "Utilizing ChatGPT to select literature for meta-analysis shows workload reduction while maintaining a similar recall level as manual curation"

### Figure 1 LARS-GPT.pdf

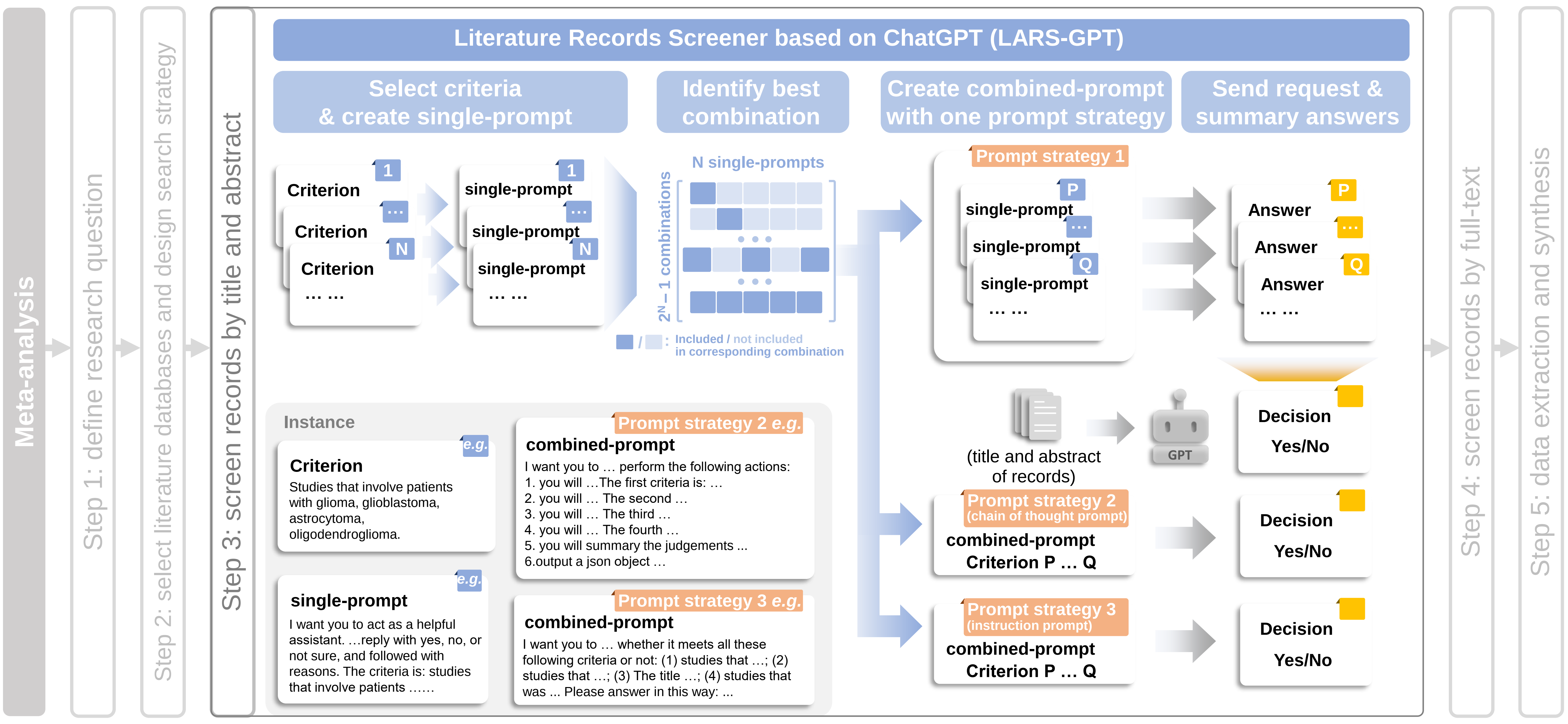

### Figure 2 Screen pipeline.pdf

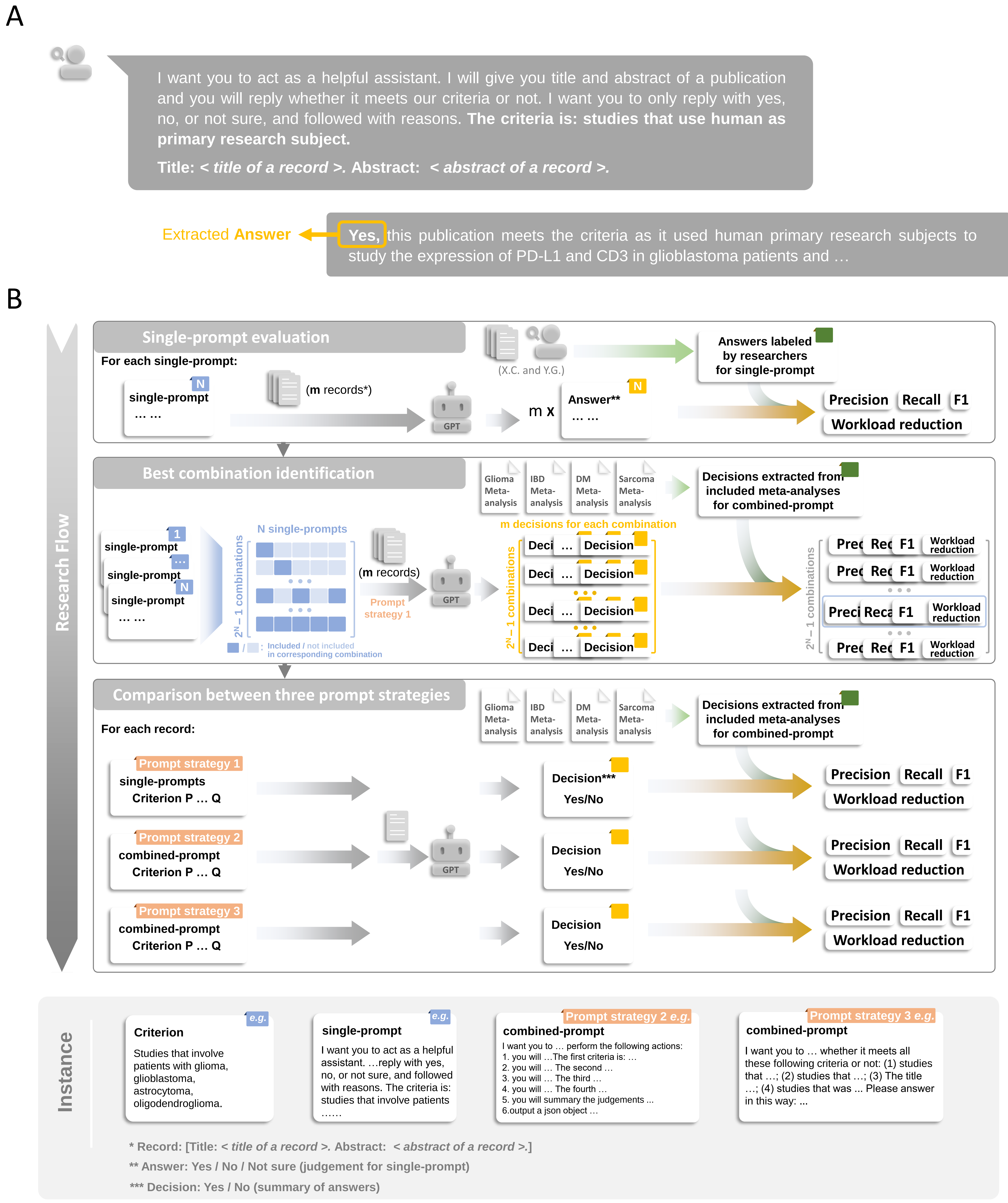

### Figure 3 prompt combination method.pdf

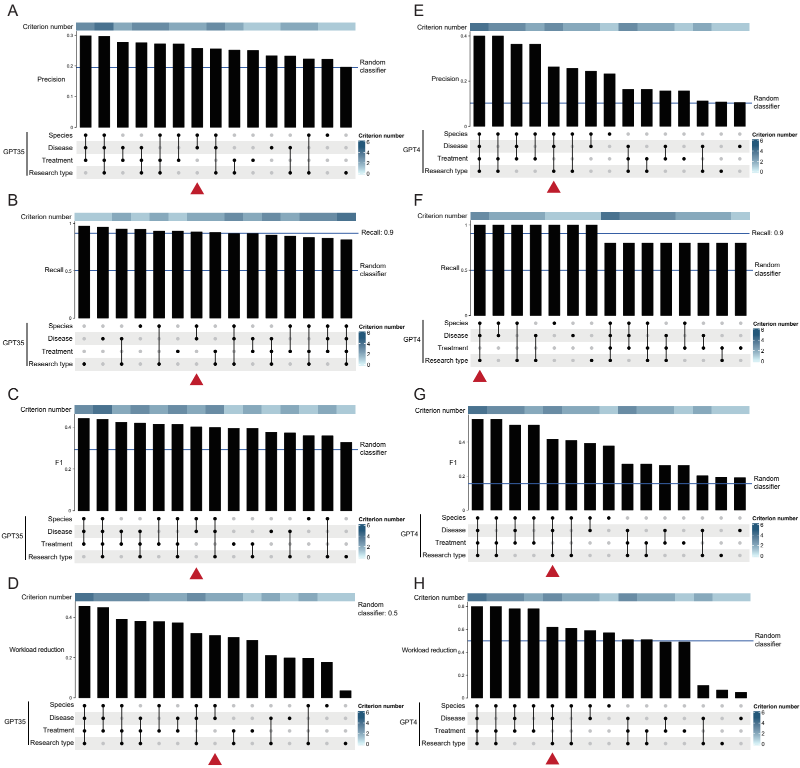
